# Preferential boosting of SARS-CoV-2 Omicron lineage-specific immune responses by monovalent XBB.1.5 vaccination

**DOI:** 10.1101/2025.10.30.25339006

**Authors:** Daryl Geers, Muriel Aguilar-Bretones, Luca M. Zaeck, Paul A. Gill, Cristina Gonzalez-Lopez, Kim Handrejk, Ngoc H. Tan, Yvette den Hartog, Lennert Gommers, Susanne Bogers, Anna Z. Mykytyn, Roos S.G. Sablerolles, Abraham Goorhuis, Douwe F. Postma, Leo G. Visser, Virgil A.S.H. Dalm, Melvin Lafeber, Neeltje A. Kootstra, Debbie van Baarle, Bart L. Haagmans, Marion P.G. Koopmans, Corine H. GeurtsvanKessel, P. Hugo M. van der Kuy, Gijsbert P. van Nierop, Menno C. van Zelm, Rory D. de Vries

## Abstract

Ongoing escape from pre-existing antibodies by severe acute respiratory distress syndrome coronavirus-2 (SARS-CoV-2) necessitates yearly coronavirus disease 2019 (COVID-19) vaccine updates for at-risk populations. Monovalent variant-specific booster vaccines, such as those for XBB.1.5, JN.1, and LP.8.1, aim to re-direct immune responses towards antigenically distinct variants; However, immune imprinting by multiple exposures to the ancestral SARS-CoV-2 spike (S) protein can hinder the de novo induction of these immune responses. Here, we profiled the SARS-CoV-2-specific immune response in healthcare workers up to 6 months after monovalent XBB.1.5 vaccination. Memory T– and B-cell responses, and antibodies were boosted up to 6 months post-vaccination. Neutralizing antibodies targeting Omicron subvariants circulating at the time of vaccination were preferentially boosted by vaccination but remained lower than those targeting the ancestral strain. Similar responses were seen for antibodies that mediate functionality through antibody-dependent cellular cytotoxicity, although these responses were more promiscuous. Broadly S-reactive B-cells were recalled with limited de novo induction of XBB.1.5-specific clones. Omicron RBD-directed B-cells targeting circulating subvariants were selected, and T-cell responses cross-reacted with all SARS-CoV-2 variants at 6 months post-vaccination. Combined, this comprehensive immune profiling demonstrates that despite evidence of imprinting, monovalent booster vaccination skews the immune response to Omicron lineage recognition.

## Introduction

The ongoing evolution of severe acute respiratory syndrome coronavirus 2 (SARS-CoV-2) gives rise to antigenically distinct variants with mutations in the spike (S) glycoprotein, facilitating evasion of pre-existing antibodies. As a response to the initial emergence of Omicron subvariants BA.1 and BA.5, bivalent vaccine boosters encoding both the ancestral S and Omicron S were introduced in late 2022. These booster vaccinations rapidly recalled immunological memory and boosted antibody and T-cell responses cross-reactive to circulating subvariants (*1*). However, ongoing antigenic drift and the emergence of novel Omicron subvariants led to reduced efficacy of these bivalent coronavirus disease-2019 (COVID-19) vaccines (*2–4*).

The World Health Organization (WHO) advises annual vaccination of at-risk groups, including the elderly, immunocompromised, and healthcare workers (HCW), with updated monovalent COVID-19 vaccines containing the most recent circulating viral variant (*5*). At the end of 2023, monovalent XBB.1.5 booster vaccines were approved and administered globally, followed by vaccines containing JN.1, KP.2, and LP.8.1 variants (*6*). Monovalent XBB.1.5 vaccination substantially boosted neutralizing antibody titers and T-cell responses against the homologous antigen, as well as against Omicron subvariants that emerged later (*7*). However, vaccine-induced neutralizing antibodies remained heavily dominated by a response towards the ancestral Wuhan-Hu-1 S, a phenomenon known as immune imprinting (*8*). This imprinting can limit the breadth of the immune response after vaccination against drifting SARS-CoV-2 variants (*9*).

Importantly, most monovalent booster immunogenicity studies have focused on neutralizing antibody measurements, overlooking other antibody effector functions, and the kinetics of B– and T-cell responses over extended periods of time. While neutralizing antibodies have been described to be predictive of immune protection (*10, 11*), these wane and can be escaped from by emerging SARS-CoV-2 variants (*12*). Consequently, just studying neutralizing antibodies provides a narrow perspective on the scope of vaccine-induced immunity. Non-neutralizing antibodies and T-cells often recognize a broader range of (conserved) epitopes within S. Non-neutralizing antibodies have been demonstrated to play a role in preventing severe COVID-19 after exposure to variants that escaped from neutralizing antibodies by targeting conserved epitopes (*13*). These antibodies are functional through the activation and degranulation of natural killer (NK) cells, opsonization of antigen for improved phagocytosis, or targeted complement deposition on infected cells (*14*). Similarly, T-cell responses are considered a correlate of protection against symptomatic and severe COVID-19 (*15*) and are known to retain cross-reactivity with emerging SARS-CoV-2 variants (*16*).

These interrelated components of the adaptive immune system are increasingly recognized for their role in protection against (severe) COVID-19 (*17, 18*). Therefore, a comprehensive immune profiling approach incorporating different components of the immune response is essential to fully understand vaccine-induced protection. To this end, we profiled the immune response of previously vaccinated HCW up to 6 months after vaccination with a monovalent XBB.1.5 booster vaccine, aiming to assess the magnitude, breadth, durability, and phenotype of humoral and cellular immune responses, and determine the impact of immune imprinting by ancestral SARS-CoV-2.

## Methods

### Study design and participants

The SWITCH-ON study is an open-label, multicenter, randomized controlled trial involving 434 healthcare workers (HCW) from four academic hospitals in the Netherlands. The study protocol (MEC-2022-0462) was approved by the Medical Ethics Committee of Erasmus University Medical Center (Rotterdam, the Netherlands), the sponsor site, and the local review boards of the other participating centers at the Amsterdam University Medical Centers, the Leiden University Medical Center, and the University Medical Center Groningen. Written informed consent was obtained from all study participants prior to the first study visit. There was no incentive or compensation for participation in the study. The study is registered with ClinicalTrials.gov (NCT05471440). The study protocol has been described before (*19*). Eligible participants had been primed with either one dose of the adenovirus-based vaccine Ad26.COV2.S (Janssen) or two doses of mRNA-based BNT162b2 (Pfizer) or mRNA-1273 (Moderna). Eligible participants were randomized to two arms: boosting with the (i) bivalent Omicron BA.1 vaccine (ancestral/BA.1) or (ii) bivalent Omicron BA.5 vaccine (ancestral/BA.5). The follow-up after bivalent vaccination has been published (*1, 20*). The focus of this study is on a sub-group of the SWITCH-ON study, in which the monovalent XBB.1.5 vaccine was offered in September 2023 to all participants. If SWITCH-ON participants planned to receive the XBB.1.5 vaccine, they were invited to join the sub-study, further referred to as the monovalent booster study (**Supplementary Figure 1**). Study samples were collected pre-vaccination (baseline; BL), at 28 post-vaccination (28D), and between 3 to 6 months post-vaccination (3/6M). Out of the 69 participants included in this study, 40 participants were randomly selected for in-depth serological profiling. Sub-analyses based on the priming COVID-19 vaccine (i.e. mRNA-based or Ad26.COV2.S-based) and bivalent booster vaccination (i.e., ancestral/BA.1 or ancestral/BA.5) were performed. Baseline characteristics of this randomly selected subgroup are described in **Supplementary Table 1**. B-cell and T-cell profiling was performed on 4 BA.1 boosted and 5 BA.5 boosted participants (9 in total) out of 40 of these individuals at BL, 28D, and 3/6M after XBB.1.5 vaccination. Due to sample availability, B-cell phenotyping was performed on a separate selection of 14 participants, which partially overlapped with the 40 participants from the sub-analyses group. At all time-points of the study, SARS-CoV-2 nucleocapsid (N)-specific antibodies were measured to establish the distribution of infections across groups. The impact of SARS-CoV-2 infection at BL or breakthrough infection throughout the study period on the kinetics of the immune response was determined separately.

### Detection of ancestral SARS-CoV-2 S1-specific binding antibodies

Serum S1-specific binding antibodies were quantified using the Liaison SARS-CoV-2 Trimeric S IgG assay (DiaSorin, #311510) as previously described (*20*). The lower limit of detection (LLoD) was set at 4.81 BAU/mL and the cut-off for positivity was set to 33.8 BAU/mL, according to the manufacturer’s instructions.

### Detection of SARS-CoV-2-binding antibodies

S-specific binding antibody levels were determined using an in-house developed ELISA assay as previously described (*20*). High-binding EIA/RIA 96-well flat-bottom plates (Corning, #3590) were coated with HEK923T cell-derived trimeric S (20 ng/mL; Sino Biological) from ancestral SARS-CoV-2 (D614G; #40589-V08H8), Omicron BA.1 (#40589-V08H26), Omicron BA.5 (#40589-V08H32), or Omicron XBB.1.5 (#40589-V08H45) at 4 °C overnight. The next day, plates were washed and blocked in Blocker BLOTTO in TBS (Fisher Scientific, #37530) supplemented with 0.01% Tween-20 at 37 °C for 1 hour. Subsequently, the plates were incubated with 4-fold serially diluted sera starting at a 1:10 dilution at 37 °C for 2 h. A PBS control and standard reference pool of 19 sera with an S1-specific binding level higher than 10,000 BAU/mL were included on every plate as controls. Following incubation, plates were washed, incubated with rabbit anti-human-IgG (1:6,000; Dako, #P0214) at 37 °C for 1 hour, washed, and developed using TMB (3,3′,5,5′-tetramethylbenzidine; SeraCare/KPL, #37530) peroxidase substrate. The reaction was stopped using an equal volume of 0.5 N sulfuric acid. Absorbance at 450 nm wavelength (OD_450_) was measured using an ELISA microtiter plate reader (Anthos 2001 microplate reader) and corrected by subtracting absorbance at 620 nm (OD_620_). A 50% endpoint titer was calculated by normalizing sample S-curves to the standard reference pool on each plate. A titer value of 10 was assigned to samples where no binding antibodies could be detected.

### Detection of SARS-CoV-2-neutralizing antibodies

Serum neutralizing antibody levels were determined by plaque reduction neutralization test (PRNT) as previously described (*20*) for seven SARS-CoV-2 variants: ancestral SARS-CoV-2 (D614G, GISAID: hCoV-19/Netherlands/ZH-EMC-2498), Delta (B.1.617.2, GISAID: hCoV-19/Netherlands/NB-MVD-CWGS2201159, Omicron BA.1 (GISAID: hCoV-19/Netherlands/LI-SQD-01032/2022), Omicron BA.5 (EVAg: 010V-04723; hCoV-19/Netherlands/ZHNEMCN5892), Omicron XBB.1.5 (GISAID: hCoV-19/Netherlands/NH-EMC-5667), Omicron BA.2.86.1 (GISAID: hCoV-19/France/IDF-IPP17625/2023), and Omicron EG.5.1 (GISAID: hCoV-19/France/GES-IPP15954/2023). Calu-3 cells (ATCC, #HTB-55) cultured in OptiMEM+GlutaMAX (Gibco, #51985-026) supplemented with 100 U/mL penicillin, 0.1 mg/mL streptomycin (Capricorn Scientific, #PS-B), and 10% fetal bovine serum (FBS; Sigma, #F7524) were used to propagate the viruses and perform the assay. In short, sera were 2-fold serially diluted in OptiMEM without FBS in different dilution ranges depending on the specific variant and S1-specific binding antibody levels to maintain the dynamic range of the assay. 400 PFU of each viral variant were incubated with the serially diluted sera at 37 °C for 1 hour. The virus-antibody mixture was transferred to Calu-3 cells and incubated at 37 °C for 8 h. Next, cells were fixed with 10% neutral-buffered formalin and permeabilized with 70% ethanol. Plaques were visualized by staining of the N protein with a polyclonal rabbit anti-SARS-CoV-2 N antibody (1:1,000; Sino Biological, #40143-T62), followed by a secondary peroxidase-labeled goat-anti-rabbit IgG antibody (1:2,000; Dako, #P0448), and color development with a precipitate-forming TMB substrate (TrueBlue; SeraCare/KPL, #5510-0030). The number of plaques per well was quantified with an ImmunoSpot Image Analyzer (CTL Europe GmbH). The 50% plaque reduction neutralization titer (PRNT50) was estimated by calculating the proportionate distance between two dilutions. An infection control (without serum) and positive serum control (Nanogam® 100 mg/mL, Sanquin) were included on every assay plate. When no neutralization was observed, the PRNT50 was assigned a value of 10.

### Detection of antibody-dependent cell-mediated cytotoxicity (ADCC)-inducing antibodies

ADCC was measured via a NK-cell degranulation assay as previously described (*21*). In short, high-binding F-bottom 96-well plates (Immunolon, Fisher Scientific, #10795026) were coated with 200 ng/well HEK293T cell-generated trimeric S (SinoBiological) from the ancestral SARS-CoV-2 (D614G; #40589-V08H8), Omicron BA.1 (#40589-V08H26), Omicron BA.5 (#40589-V08H32), or Omicron XBB.1.5 (#40589-V08H45) at 4 °C overnight. The antigens are the same as the ones used for the ELISA. Plates were blocked, washed, and incubated with 4-fold serially diluted sera starting at a 1:40 dilution at 37 °C for 2 h. A PBS control and standard reference pool consisting of 19 sera with an S1-specific binding level higher than 10,000 BAU/mL as used for the ELISA were included on every plate. Following serum incubation, plates were washed and 100,000 NK92.05 cells expressing the CD16 Fc-receptor were added supplemented with CD107a^V450^ (1:100, clone H4A3, BD, #561345), GolgiStop (0.67 mL/mL, BD, #554724), and GolgiPlug (1 mL/mL, BD, #555029). Plates were incubated at 37 °C for 5 h, washed and stained for viable NK cells with CD56^PE^ (1:25, clone B159, BD, #555516) and LIVE/DEAD Fixable Aqua Dead Cell (AmCyan, Invitrogen, 1:100, #L34957). Cells were stained at 4 °C for 30 minutes and fixed in Cytofix (BD, #554722) at 4 °C for 30 minutes. NK92.05-CD16 cells were measured using a FACSLyric (BD) and degranulating cells were identified as CD56+CD107a+ cells. S-curves were generated by using the lowest and highest percentage of degranulation normalized to the reference pool, from which a 50% endpoint titer was calculated. A titer value of 10 was assigned to samples where no binding antibodies could be detected.

### Detection of recent SARS-CoV-2 infections

Infections were identified via the detection of SARS-CoV-2 N-specific antibodies in serum. N-specific antibodies were measured at BL, 28D, and 3/6M after monovalent XBB.1.5 vaccination using a SARS-CoV-2 IgG kit (Abbott, #06R-86-22) according to manufacturer’s instructions. N-specific antibody levels were reported as a signal-to-cut-off (S/CO) ratio. A positivity cut-off was set at an S/CO ratio of ≥1.4, according to manufacturer’s recommendations. N-positive participants were not excluded from further analysis. Instead, a separate analysis on S-specific antibody kinetics was performed for participants with detectable N-specific antibodies at BL and for participants with a breakthrough infection (BTI) between BL and 28D or between 28D and 3/6M after monovalent XBB.1.5 vaccination (**Supplementary Figure 2**).

### B-cell culture and differentiation into antibody-secreting cells

To determine B-cell cross-reactivity patterns, peripheral-blood-derived B-cells were isolated and stimulated, expanded, and differentiated into antibody secreting cells (ASC) *in vitro*. Oligoclonal B-cell cultures were performed at limiting density to ensure the presence of a single SARS-CoV-2-reactive B-cell per well, as described before (*22*). In brief, CD19+ B-cells were isolated from PBMC using magnetic EasySep™ Human CD19 Positive Selection Kit II (STEMCELL technologies, #17854). Oligoclonal B-cell cultures of 50 to 100 CD19+ B-cells were co-cultured with 10 times the number of irradiated CD40L-expressing fibroblasts in AIM-V AlbuMAX culture medium (Thermo Fisher #31035025) supplemented with 10% FBS, 100 U/mL penicillin, 0.1 mg/mL streptomycin (Capricorn Scientific, #PS-B), and 50mM β-mercaptoethanol (Gibco, #11528926) (B-cell medium). CD40L-expressing fibroblasts were provided by J. Banchereau from The Jackson Laboratory for Genomic Medicine, Farmington, Connecticut, USA. Co-cultures of B-cells and CD40L-expressing fibroblast were initiated in B-cell medium supplemented with 50 U/ml IL-2 (Novartis), 10 ng/ml IL-10 (Peprotech, #AF-200-10), 1 µg/ml Resquimod (InvivoGen, #tlrl-r848-1) and 25 ng/ml IL-21 (Peprotech, #200-21) for 72 h. Subsequently, culture supernatants were replaced with new B-cell medium supplemented with 25 ng/mL IL-21 (PeproTech, #200-21). ASC culture supernatants were harvested 14 days after initiation and stored at –20 °C for profiling of IgG cross-reactivity patterns.

### Profiling of IgG cross-reactivity patterns

To determine the breadth of the B-cell response, IgG reactivity towards an array of SARS-CoV-2 antigens was screened in ASC culture supernatants using protein microarray (PMA) as described elsewhere (*23*). PMA slides contained the pre-fusion stabilized S and receptor-binding domain (RBD, all Sino Biological) of ancestral SARS-CoV-2 (#40589-V08H8 and #40592-V08H), Delta (#40589-V08H10 and #40592-V08H90), Omicron BA.1 (#40589-V08H26 and #40592-V08H121), BA.5 (#40589-V08H32 and #40592-V08H130), B.Q.1.1 (#40589-V08H41 and #40592-V08H143), BA.2.86 (#40589-V08H58 and #40592-V08H152), EG.5.1 (#40589-V08H55 and #40592-V08H151), JN.1 (#40589-V08H59 and #40592-V08H155), and XBB.1.5 (#40589-V08H45 and #40592-V08H146). All antigens were titrated and the minimal amount that gave saturated fluorescent signal using highly reactive control sera was determined. All antigens were spotted in equimolar amounts (0.345 ng – 0.460 ng per antigen and spot) in duplicate on nitrocellulose pads (64-pad glass slides; Sartorius) using a sciFlexArrayer SX (Scienion). PMA slides were first incubated with Blocker™ BLOTTO in TBS (Blocking Buffer, BB, #37530) for 1 hour at 37 °C and then washed with PBS + 0.05% Tween-20 (Washing Buffer, WB). Subsequently, a 1:1 mix of BB with individual culture supernatants was incubated for 1 hour at 37 °C on each pad. Slides were then washed 5 times with WB and incubated with BB containing goat polyclonal anti-human IgG conjugated with Alexa Fluor 647 (1:500, Jackson ImmunoResearch, #109-605-098) for 1 hour at 37 °C. PMA slides were washed 5 times with WB and once with double-distilled water, air dried and scanned on a PowerScanner (Tecan). The mean fluorescence intensity (MFI) of IgG spots on PMA slides was analyzed using ScanArray Express software. MFI values of duplicate antigens for every nitrocellulose pad were averaged. Culture supernatants were regarded as reactive for that antigen when the signal for that antigen was above 1,000 MFI (detection range 0 – 65,000 MFI) based on background reactivity in negative protein array pads.

### Immunophenotyping of SARS-CoV-2 RBD-specific memory B cells

Fluorescent tetramers were made from RBD proteins of ancestral SARS-CoV-2, Omicron BA.1, BA.5, XBB.1.5 and JN.1, and generated as previously described (*24*). In short, DNA constructs were cloned into a pCR3 plasmid and produced using the Expi293 Expression system (Thermo Fisher, Waltham, MA), purified, biotinylated, and tetramerized. The following fluorescent tetramers were generated: [RBD ANC]_4_^BUV395^, [RBD ANC]_4_^BV421^, [RBD XBB.1.5]_4_^BV480^, [RBD XBB.1.5]_4_^BUV737^, [RBD BA.1]_4_^BUV496^, [RBD BA.5]_4_^BUV615^, and [RBD JN.1]_4_^BV650^. These tetramers were incorporated into a 20-color spectral flow cytometry panel for B-cell phenotyping (**Supplementary Table 2**). 5–15 × 10^6^ thawed PBMC were incubated at room temperature in the dark for 15 min in a total volume of 250 µL with FACS buffer (PBS with 0.1% sodium azide and 0.2% BSA; FB). Next, PBMC were stained in 100 µL with the following antibodies/dyes in their respective dilutions: fixable ViaDye Red LIVE/DEAD marker (1:4000), CD3^BUV805^ (UCHT1, 1:40), CD19^BYG710^ (HIB19, 1:100), CD21^BV711^ (B-ly4, 1:20), CD27^BYG781^ (O323, 1:200), CD38^BV605^ (HB-7, 1:500), CD71^BV786^ (M-A712, 1:100), IgM^V450^ (MHM88, 1:400), IgD^BB700^ (IA6-2, 1:100), IgG1^PE^ (G17-1, 1:1000), IgG2^FITC^ (HP6002, 1:100), IgG3^FITC^ (HP6047, 1:200), IgG4^APC^ (SAG4, 1:50), IgA^PE-Vio615^ (RE1014, 1:66), and 5 µg/mL of each in-house generated RBD fluorescent tetramer. In a separate tube, 1–5 × 10^6^ PBMC were incubated in parallel in a total volume of 100 µL with FB containing the following antibodies/dyes and their respective dilutions: fixable ViaDye Red LIVE/DEAD marker (1:4000), CD3^BUV805^, CD19^BYG710^, CD27^BYG781^, and IgD^BB700^ and fluorochrome-conjugated streptavidin controls (BUV395, BV421, BV480, BUV737, BUV496, BUV615 and BV650) without respective RBD proteins (**Supplementary Table 2**). Following staining, cells were washed, fixed with 2% PFA for 20 min at room temperature in the dark, and washed once more before acquisition on a 5-laser Cytek Aurora (Cytek Biosciences) using SpectroFlo® software v3.3.0. Data analysis was performed using FlowJo™ Software v10.10.0.

### Detection of SARS-CoV-2-specific T-cell responses by interferon gamma release assay

T-cell response profiles were generated from samples obtained 6 months after monovalent XBB.1.5 vaccination as previously described for earlier timepoints post vaccination (*16*). Interferon gamma (IFNγ) responses were measured after stimulation of whole blood with peptide pools covering the S protein of ancestral SARS-CoV-2 (#PM-WCPV-S-1), Delta B.1.617.2 (#PM-SARS2-SMUT06-1), Omicron BA.1 (#PM-SARS2-SMUT08-1), BA.2 (#PM-SARS2-SMUT09-1), BA.5 (#PM-SARS2-SMUT10-1), BQ.1.1 (#PM-SARS2-SMUT13-1), XBB.1.5 (#PM-SARS2-SMUT15-1), and EG.5.1 (#PM-SARS2-SMUT16-1), and hCoV-OC43 (#PM-OC43-S-2), hCoV-NL63 (#PM-NL63-S-1), and MERS-CoV (#PM-MERS-CoV-S-1) S (1 mg/mL; JPT technologies). In short, peptide pools were added to 200 µL lithium heparin (LiHep) anticoagulated whole blood and incubated in 1.4 mL polypropylene tubes in a 96-tube rack (Micronic) at 37 °C for 18 h. As a negative control, LiHep whole blood was incubated in negative control (NIL) tubes from the QuantiFERON test (Qiagen). A super stimulation (CEFX, 1 mg/mL, JPT technologies, #PM-CEFX-1) pool containing commensals and common pathogens was used as a positive control. Following incubation, whole blood was transferred to a 96-well V-bottom plate and centrifuged at 2,500xg for 15 minutes. Next, plasma was collected, and IFNγ levels were measured using the QuantiFERON SARS-CoV-2 ELISA kit (Qiagen). In short, 50 µL plasma was incubated for 2 h with enzyme-conjugate solution (1:1 ratio) on the pre-coated ELISA plates. These were washed 4 times with wash buffer, developed for 10 minutes using substrate solution, and the color development was terminated using a stop solution included in the ELISA kit. Absorbance at 450 nm wavelength (OD_450_) was measured using an ELISA microtiter plate reader (Anthos 2001 microplate reader) and corrected by subtracting absorbance at 620 nm (OD_620_). IFNγ levels were calculated from a standard curve with known IFNγ concentrations, corrected by subtracting the NIL value, and depicted as IU/mL. A 0.15 IU/mL responder cut-off was maintained as previously described and per manufacturer’s instructions. The LLoD was set at 0.01 IU/mL as per manufacturer’s instructions and undetectable IFNγ levels were assigned this value.

### Phenotyping SARS-CoV-2-specific T-cells

Immune phenotyping of major immune cell subsets in peripheral blood was performed using CyTOF (cytometry by time of flight; Fluidigm). PBMC were thawed and washed once in ice-cold Iscove′s Modified Dulbecco′s Medium (IMDM, #12440053) supplemented with 10% fetal bovine serum, 100 U/mL penicillin, and 0.1 mg/mL streptomycin (Capricorn Scientific, #PS-B). Thereafter, cells were incubated with 10,000 IU/mL Benzonase (Merck, #70664-3) for 30 minutes at 37 °C, washed, and resuspended in Roswell Park Memorial Institute (RPMI, Invitrogen, #21875091) supplemented with 10% human pooled serum, 100 U/mL penicillin, and 0.1 mg/mL streptomycin (Capricorn Scientific, #PS-B). Next, PBMC were seeded in U-bottom 96-well plates and rested at 37 °C overnight. Consecutively, PBMC were stimulated with overlapping peptide pools covering the ancestral SARS-CoV-2 (#PM-WCPV-S-1) or Omicron XBB.1.5 (#PM-SARS2-SMUT15-1) S (1 µg/mL, JPT technologies) and incubated at 37°C for 20 h. PBMC were stimulated with an equimolar concentration of DMSO or a super stimulation (CEFX, 1 μg/mL, JPT technologies, #PM-CEFX-1) as a negative and positive control, respectively. Following stimulation, PBMC were washed with Maxpar Cell Staining Buffer (Standard Biotools, #201068) and barcoded in the residual volume by adding 0.5 µL CD45^113Cd^ and CD45^194Pt^ (ancestral S), CD45^194Pt^ and CD45^195Pt^ (XBB.1.5 S), or CD45^113Cd^ and CD45^195Pt^ (DMSO; **Supplementary Table 3**). After 15 minutes at RT, cells were washed twice with cell staining buffer, and barcoded cells of the same donor were pooled and resuspended in 270 µL cell staining buffer containing 3 µL drop-in antibodies for T-cell phenotyping antibodies (**Supplementary Table 3**). Cells were further stained using the Maxpar Direct Immune Profiling Assay Cell (MDIPA) staining kit (Standard Biotools, #201334; **Supplementary Table 3**) according to the manufacturer’s instructions. In brief, cells in 270 µL were added to an MDIPA tube containing a lyophilized immune phenotyping antibody panel and incubated for 30 minutes at RT. Subsequently, cells were fixed with 1.6 % formaldehyde, stained with 31.25nM iridium DNA intercalator (part of the staining kit) and stored at –80 °C until further use. Prior to acquisition, samples were thawed, washed twice with cell staining buffer and cell acquisition solution, and resuspended at 1 × 10^6^ cells per ml in cell acquisition solution containing 1:10 diluted EQ normalization beads (Fluidigm, #201078). Samples were measured on a Helios Mass Cytometer at an event rate below 300 events per second.

Measurements were normalized using the Fluidigm software 7.0.8493 and cleaned using the Pathsetter software v.3. Using the OMIQ software (Dotmatics), cleaned data files were initially used for identifying AIM+ T-cells and further immune cell phenotyping was performed by unsupervised cluster analysis using FlowSOM. For classical gating, LIVE CD3+ T-cells were gated for CD4+ memory, or CD8+ memory cells and AIM+ cells were identified as CD4+CD137+CD69+ or CD8+CD137+CD69+ T-cells, respectively. For phenotyping, FCS files were concatenated, scaled, and normalized to 60,000 live CD3+ events per sample to run a opt-SNE using the following markers: CD196, CD4, CD8, CD45RO, CD45RA, CD194, CD25, CD27, CD57, CD183, CD185, TIGIT, CD28, CD38, TCRγδ, CD279, CD294, CD197, CD366, HLA-DR, CD278, and CD127. Next, a SOM (self-organized map) was created with 15 clusters and conventionally gated AIM+ T-cells were back-gated onto opt-SNE to phenotype antigen-specific T-cell responses.

### Statistical analysis

This analysis represents a sub-study of a larger clinical trial (*25*) This sub-study was not pre-specified in the trial protocol; therefore no power calculation was conducted. The analyses were conducted on the available cohort and are observational and exploratory in nature.

Baseline characteristics were summarized descriptively. Continuous variables with non-normal distributions were reported as medians with interquartile ranges (IQR), and categorical variables as counts and percentages. Missing values were not imputed. Immunological outcomes were expressed as geometric mean titers (GMT) or geometric means (GM) with 95% confidence intervals (CI). Between-group comparisons of unpaired, non-parametric data (e.g., priming or bivalent vaccination groups) were assessed using the Mann–Whitney U test. To account for multiple comparisons, Bonferroni correction was applied for 4 independent comparisons. Therefore, *P*-values of less than 0.01 were used as the statistical significance threshold. Statistical significance of paired B-cell phenotyping data was assessed using the Wilcoxon matched-pairs signed rank test. Correlation matrices of T-cell responses across variants were constructed by calculating Pearson correlation coefficients. All figures were generated using GraphPad Prism (version 10.6.0) and finalized in Adobe Illustrator 2025.

## Results

### Baseline characteristics

A total of 86 HCW were enrolled in the XBB.1.5 monovalent booster study (*19*). Participants for whom samples at baseline or on day 28 after XBB.1.5 monovalent booster vaccination were not available were excluded (n=16). An additional participant (n=1) was excluded that did not receive bivalent booster vaccination, resulting in a total of 17 exclusions. Thus, 69 HCW were included for analyses of which baseline characteristics are described in **Table 1**. The number of female participants was higher than the number of male participants per subgroup and overall; this reflects the current female to male ratio of the healthcare workforce (*26*). The median age across all sub-groups was similar (range of medians 53 to 58 years). The median time between bivalent BA.1 bivalent and monovalent booster was higher than between bivalent BA.5 and monovalent booster, reflecting the later application of bivalent BA.5 in the Dutch vaccination campaign. Notably, participants who had been primed with Ad26.COV2.S had lower S1-binding antibody levels at baseline than participants who had received an mRNA-based vaccine as their priming vaccine (**Table 1**). This is in line with earlier observations in the SWITCH and SWITCH-ON study (*20*).

**Table 1.** Baseline characteristics.

|  | Total<br>n = 69 | Ad/BA.1<br>n = 20 | mRNA/BA.1<br>n = 29 | Ad/BA.5<br>n = 12 | mRNA/BA.5<br>n = 8 |
| --- | --- | --- | --- | --- | --- |
| **Demographics** |  |  |  |  |  |
| Female [n (%)] | 47 (68%) | 13 (65%) | 22 (76%) | 5 (42%) | 7 (88%) |
| Age, years [median (IQR)] | 55 (50-58) | 56 (50-58) | 53 (45-56) | 56 (52-58) | 58 (54-64) |
| **Comorbidities** |  |  |  |  |  |
| Cardiovascular disease [n (%)] | 2 (2%) | 0 (0%) | 1 (3%) | 1 (8%) | 0 (0%) |
| Pulmonary disease [n (%)] | 3 (4%) | 0 (0%) | 2 (7%) | 1 (8%) | 0 (0%) |
| Diabetes Mellitus [n (%)] | 1 (1%) | 1 (5%) | 0 (0%) | 0 (0%) | 0 (0%) |
| Liver disease [n (%)] | 1 (1%) | 0 (0%) | 0 (0%) | 1 (8%) | 0 (0%) |
| Kidney disease [n (%)] | 1 (1%) | 1 (5%) | 0 (0%) | 0 (0%) | 0 (0%) |
| **Immunology** |  |  |  |  |  |
| S1-binding antibodies,<br>BAU/mL [GMT (95% CI)] | 2,834<br>(2,221-3,615) | 1,962<br>(1,180-3,260) | 4,338<br>(3,090-6,090) | 1,603<br>(1,159-2,217) | 3,445<br>(1,269-9,354) |
| **Time between visits** |  |  |  |  |  |
| Last vaccination and BL, days<br>[median (IQR)] | 377<br>(348-397) | 377<br>(371-385) | 394<br>(381-406) | 310<br>(301-330) | 339.0<br>(320-360) |
| BL and 28D, days<br>[median (IQR)] | 28<br>(27-28) | 28<br>(27-28) | 28<br>(27-28) | 28<br>(27-28) | 28<br>(27-29) |
| 28D and 3/6M, days<br>[median (IQR)] | 133<br>(71-176) | 135<br>(128-143) | 118<br>(105-130) | 209<br>(186-219) | 186<br>(170-196) |

BAU: binding arbitrary units; GMT geometrical mean titer; 28D: 28 days, and 3/6M: 3 to 6 month after XBB.1.5 vaccination; BL: base line; IQR: interquartile range

### Monovalent XBB.1.5 vaccination increases antibody levels

To evaluate the immunogenicity of the monovalent XBB.1.5 vaccine, ancestral SARS-CoV-2-specific binding and functional antibody responses were measured up to 6-month post-vaccination (**Figure 1**). Monovalent XBB.1.5 booster vaccination increased S1-specific and trimeric S IgG binding antibodies across all groups, with peak antibody levels at day 28 post-vaccination (**Figure 1A and B**) followed by waning at 3-6 months post-vaccination, which did not fully return to baseline. No significant differences were observed between the bivalent ancestral/BA.1 or ancestral/BA.5 vaccine groups at any timepoint (*P*=0.12 [BL], *P*=0.16 [28D], and *P*=0.04 [3/6M]). However, mRNA-primed individuals had consistently higher S1-specific binding antibody levels (*P*=0.0006 [BL], *P*<0.0001 [28D], and *P*=0.0003 [3/6M]) than those primed with Ad26.COV2.S, independent of bivalent vaccination history. Interestingly, no significant differences were found between mRNA-and Ad26.COV2.S-primed individuals when assessing trimeric S antibody levels (**Figure 1B**). Similarly, ancestral-specific neutralizing and ADCC-mediating antibody levels increased 28 days after vaccination (**Figure 1C and D**). While neutralizing antibody levels increased equally in all subgroups, ADCC-mediating antibody levels were higher in Ad26.COV2.S-primed individuals (**Figure 1D**). N-specific antibody analyses revealed a large proportion (27/69; 39%) of SARS-CoV-2 infections before monovalent XBB.1.5 vaccination (baseline). Within N-negative participants (n=42), 2 breakthrough infections occurred between BL and 28D (5%), and 13 breakthrough infections between 28D and 3/6M (22%; **Supplementary Figure 2A**). However, neither a recent infection before vaccination (**Supplementary Figure 2B**) nor breakthrough infections (**Supplementary Figure 2C**) affected the rate of antibody waning.

**Figure 1.**
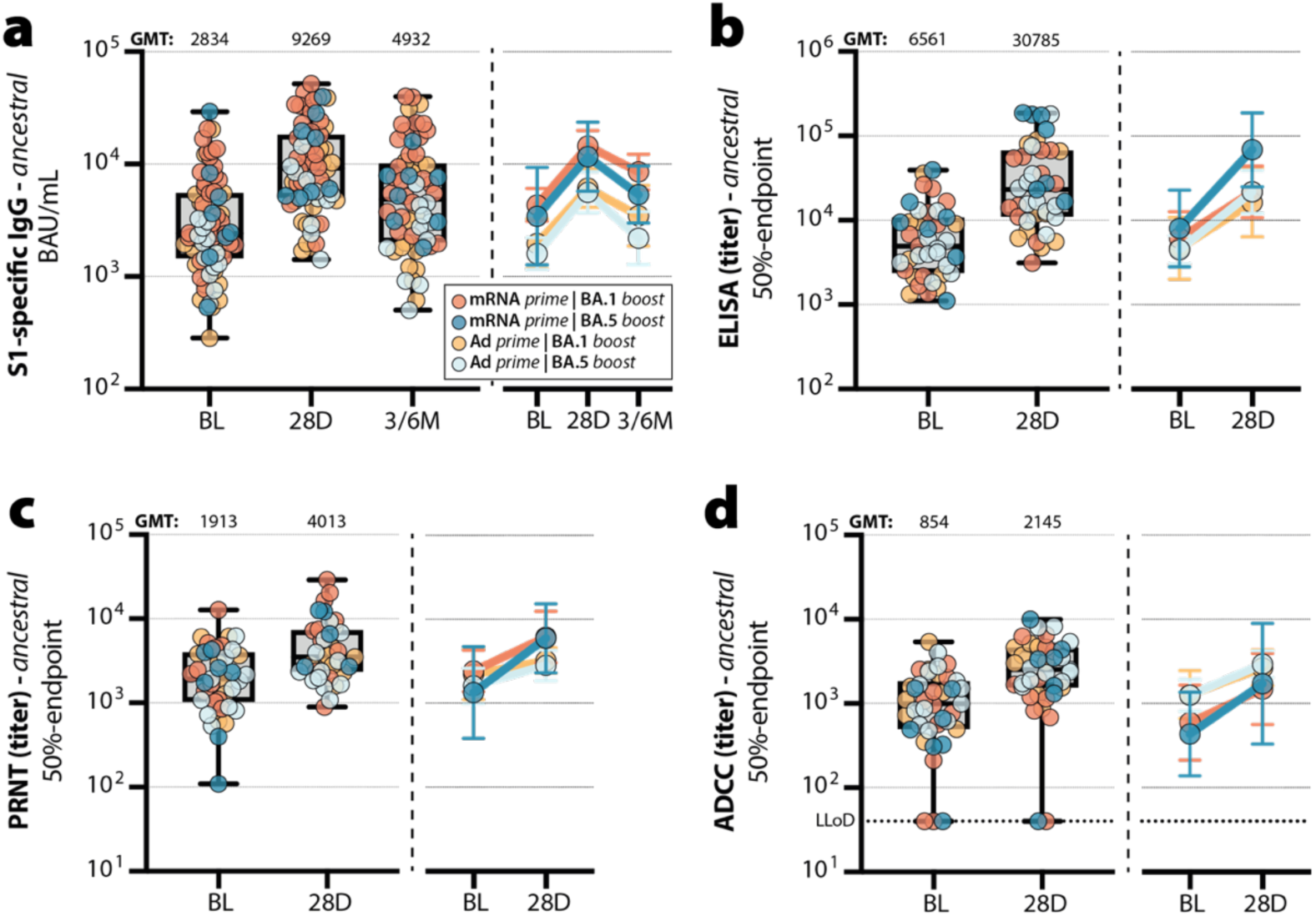
Ancestral-specific antibodies after monovalent XBB.1.5 vaccination. **(A)** S1-specific IgG binding antibody levels, **(B)** trimeric S-specific IgG binding antibody levels, **(C)** neutralizing antibody levels (measured by PRNT), and **(D)** ADCC-mediating antibody levels pre– and post XBB.1.5 vaccination. Colors represent different prime-boost regimes: mRNA prime, BA.1/anc bivalent boost (orange); mRNA prime, BA.5/anc bivalent boost (blue), Ad26.COV2.S prime, BA.1/anc bivalent boost (yellow); and Ad26.COV2.S prime, BA.5/anc bivalent boost (light blue). The horizontal lines of the box-and-whisker plots indicate the median, the whiskers indicate the range, and the bounds of the boxes indicate the interquartile range (IQR). All individual datapoints are shown in the box-and-whisker plots. Geometric means are depicted on top of each box. Line graphs next to each panel depict the geometric means with 95% confidence intervals for each prime-boost group. Mann-Whitney test for non-parametric unpaired comparisons between groups was performed to test significance.

### Breadth of neutralizing antibodies is increased by XBB.1.5 vaccination

To further characterize the breadth of vaccine-induced antibody responses, we assessed binding and functional antibody breadth across ancestral SARS-CoV-2, Delta and Omicron subvariants following monovalent XBB.1.5 vaccination. In all cases, the absolute levels were assessed, followed by fold increases compared to the baseline level, and relative levels at day 28 post-vaccination in comparison to the ancestral SARS-CoV-2 response (**Figure 2**). Monovalent XBB.1.5 vaccination boosted trimeric S-reactive antibodies to all tested variants at 28 days post-vaccination, with the highest titers measured against ancestral S (**Figure 2A**). However, a preferential increase for antibodies binding to Omicron variants, particularly XBB.1.5, was observed (**Figure 2B**). Despite this preferential boost for Omicron XBB.1.5, binding antibody levels to Omicron subvariants remained considerably lower than to ancestral SARS-CoV-2 (55% relative to ancestral) (**Supplementary Figure 3A**). Similarly, neutralizing antibodies were boosted post-vaccination but remained dominated by responses to ancestral and Delta SARS-CoV-2, with lower reactivity against Omicron subvariants, including XBB.1.5, BA.2.86, and EG.5.1 (**Figure 2C**). A notable boost of neutralizing antibodies against Omicron variants XBB.1.5 (5.9-fold increase), BA.2.86 (5.2-fold increase), and EG.5.1 (6.9-fold increase) was observed (**Figure 2D**). However, a narrow reactivity pattern dominated by the ancestral strain was observed across all prime-boost groups, with notably less neutralization detected against the XBB.1.5 Omicron variant (**Supplementary Figure 3B**). The trends for ADCC-mediating antibodies were similar to those observed with trimeric S-specific binding antibodies (**Figure 2E**). Although ADCC-mediating responses were 2-fold lower for the XBB.1.5 variant as compared to ancestral SARS-CoV-2 **(Figure 2E**), a preferential boost for XBB.1.5 was observed (**Figure 2F**). Notably, ADCC-mediating antibodies retained a higher degree of cross-reactivity between Omicron subvariants and ancestral SARS-CoV-2 (up to 70% relative to ancestral; **Supplementary Figure 3C**).

**Figure 2.**
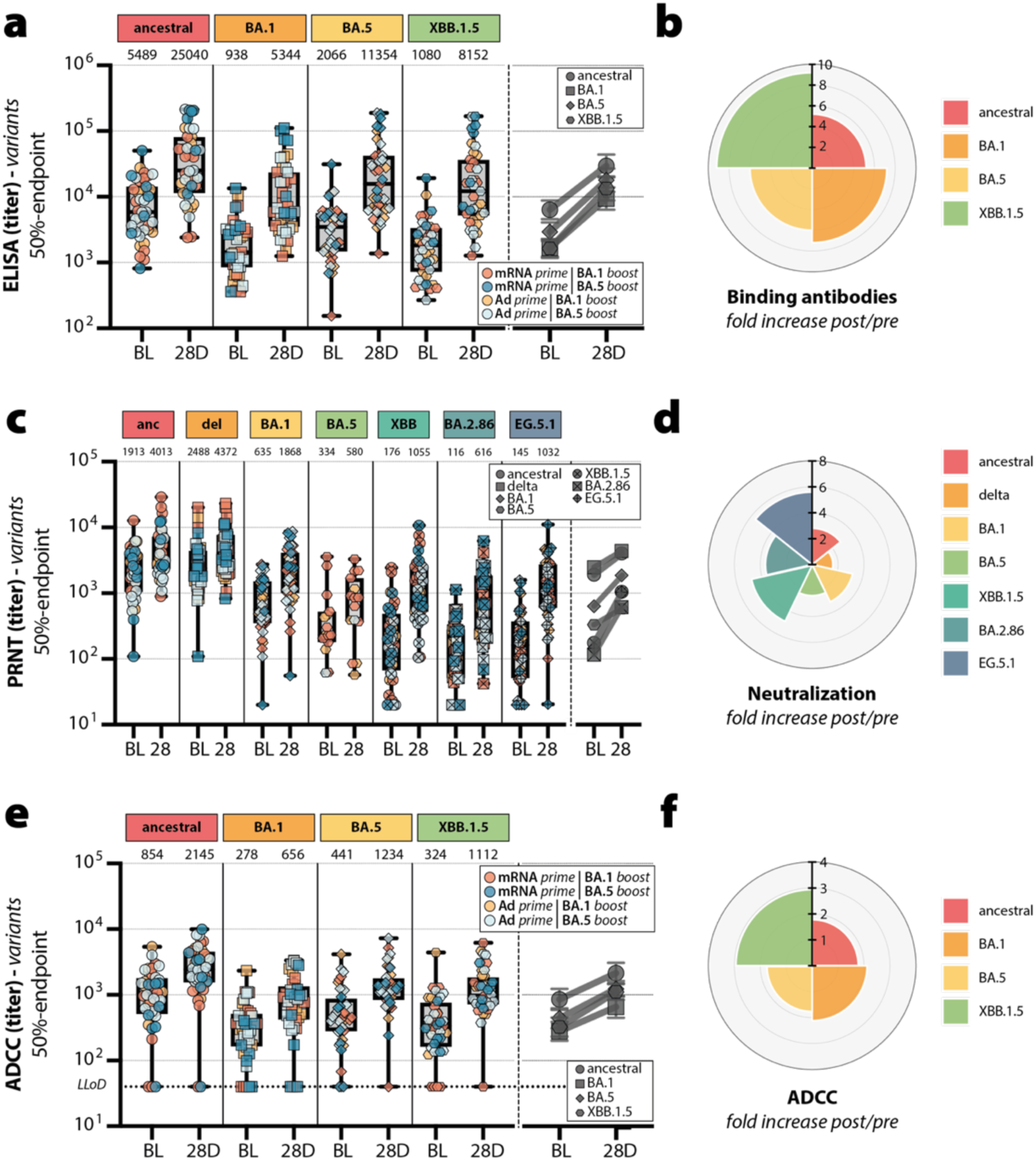
Breadth of neutralizing antibody response after monovalent XBB.1.5 vaccination. **(A)** Trimeric S-specific binding antibody levels for ancestral SARS-CoV-2, Omicron BA.1, BA.5, and XBB.1.5 subvariants. **(B)** Fold increase in variant-specific binding antibody levels from baseline to 28 days after XBB.1.5 vaccination. **(C)** Neutralizing antibody levels against ancestral SARS-CoV-2, Delta, Omicron BA.1, BA.5, XBB.1.5, BA.2.86, and EG.5.1 subvariants. **(D)** Fold-increase in variant-specific neutralizing antibody levels from baseline to 28 days after XBB.1.5 vaccination. **(E)** ADCC-mediating antibody levels for ancestral SARS-CoV-2, Omicron BA.1, BA.5, and XBB.1.5 subvariants. **(F)** Fold increase in variant-specific ADCC-mediating antibody levels from baseline to 28 days after XBB.1.5 vaccination. Colors represent different prime-boost regimes and SARS-CoV-2 variants are indicated by different symbols. The horizontal lines of the box-and-whisker plots indicate the median, the whiskers indicate the range, and the bounds of the boxes indicate the interquartile range. All individual datapoints are shown in the box– and-whisker plots. Geometric mean titers are depicted on top of each box. Line graphs next to each panel depict the geometric means with 95% confidence intervals for each SARS-CoV-2 variant over time.

### Monoclonal antibody cross-reactivity is broadened after monovalent XBB.1.5 vaccination

To determine changes in frequency and unravel variant-specificity or cross-reactivity of circulating IgG B-cell clones before and after monovalent XBB.1.5 vaccination, the clonal reactivity of antibodies derived from CD19+ B-cells was assessed in 9 individuals up 6 months post-vaccination. S trimers and RBD of ancestral SARS-CoV-2, Delta, and various Omicron subvariants (BA.1, BA.5, BQ.1.1, XBB.1.5, EG.5.1, BA.2.86) were considered at all timepoints; Omicron JN.1 was included at 3-6 months post vaccination.

On average, high frequencies of IgG B-cell clones with reactivity to all variant S were detected in circulation prior to vaccination (**Figure 3A and Supplementary Figure 4A**). Nevertheless, frequencies of reactive IgG B-cell clones increased 28 days after vaccination. Their numbers uniformly reduced at 3-6 months after vaccination to levels below those found at baseline. Similar to S, the frequency of RBD-reactive IgG B-cell clones was boosted after monovalent XBB.1.5 vaccination for all variants (**Figure 3A**). Notably, RBD-reactive clones were boosted significantly more and waned less compared to S-reactive clones, and with more variation between variants (**Figure 3B and Supplementary Figure 4B**). Consequently, while RBD-reactive clones targeting ancestral SARS-CoV-2, Delta and BA.1 showed a net decrease at 3-6 months compared to baseline, BA.5, BQ.1.1, XBB.1.5, EG.5.1 and BA.2.86 showed a net increase (**Figure 3B**).

**Figure 3.**
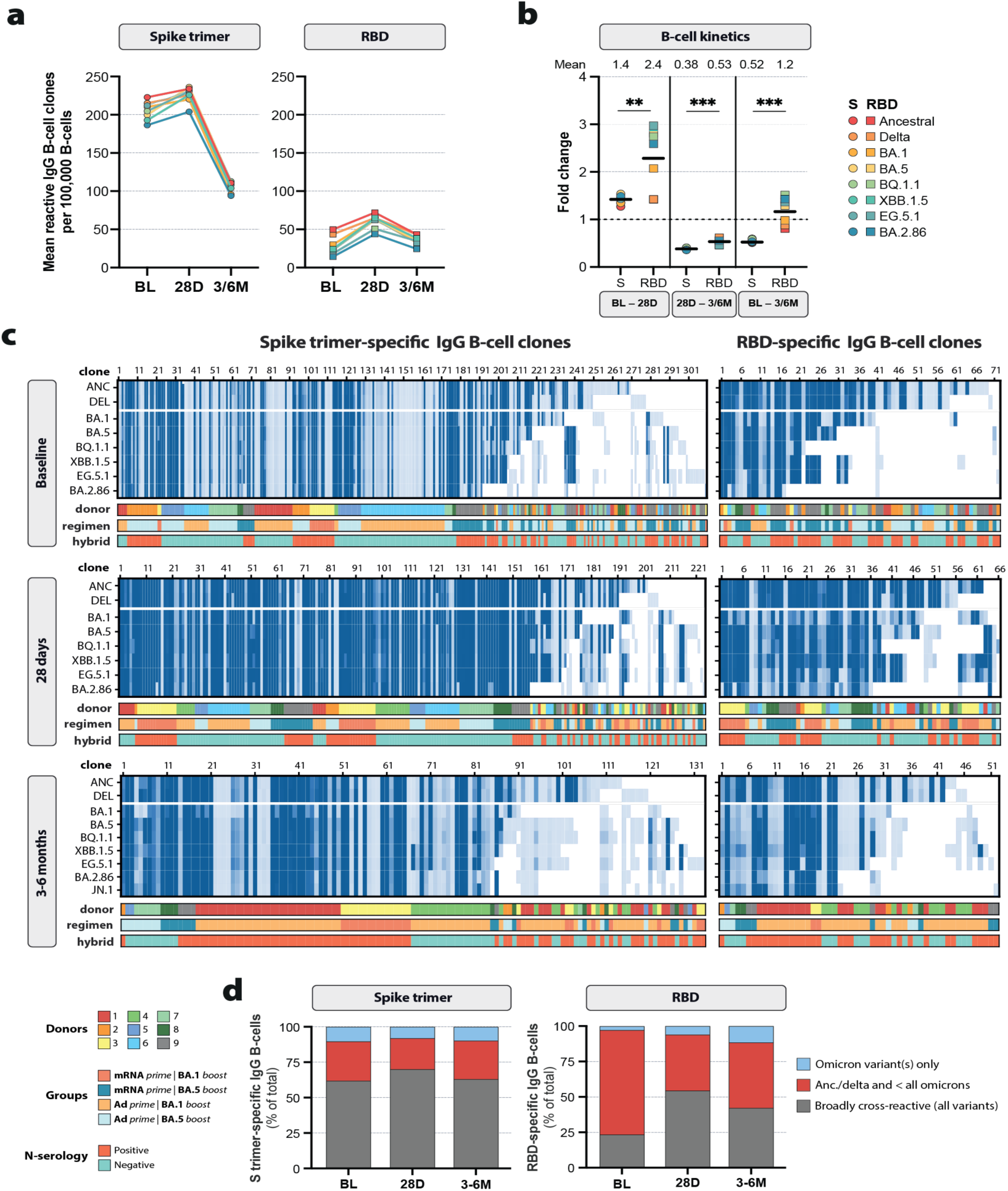
B-cell profiling and cross-reactivity analysis. **(A)** Mean of SARS-CoV-2-reactive B-cells per 100,000 PMA-screened B-cells targeting the S trimer and RBD at baseline (BL), 28 days (28D) and 3/6 months (3/6M) post-XBB.1.5 booster, color coded per variant, assessed using a PMA. **(B)** Mean fold changes between baseline (BL), 28 days (28D) and 3/6 months (3/6M) after booster for Spike (S) and RBD-specific B-cell frequencies, color coded per variant. The mean fold change for all variants is indicated with a horizontal line and specified above the graph. **(C)** IgG cross-reactivity patterns towards S trimer and RBD for the indicated SARS-CoV-2 variants were generated for each time point. Clonal IgG reactivity of culture supernatants was assessed using a PMA. Light to dark shades of blue represent the mean fluorescent intensity (reactivity) towards each antigen (light blue 1,000 to dark blue 65,000). Clones are color-coded based on their donor number, history of prime-boost regimen, and detection of a breakthrough infection. **(D)** Fraction of IgG clones that cross-recognize all variant S trimers and RBD (gray), Ancestral/Delta variants and not all omicrons (red), or target Omicron variants only (light blue).

Prior to vaccination, a total of 311 SARS-CoV-2 S-specific IgG B-cell clones were detected and analyzed for their (cross-)reactivity across all 9 individuals (**Figure 3C, left panels**). Most IgG B-cell clones were broadly cross-reactive to ancestral SARS-CoV-2, Delta and all assessed Omicron subvariants up to BA.2.86 S (clones 1-192 [62%]; **Figure 3D and Supplementary Figure 4C**). Ancestral S-reactive IgG B-cell clones dominated the SARS-CoV-2-specific B-cell response in all individuals, followed by Delta-reactive clones. At 28 days after XBB.1.5 vaccination, 224 S-reactive IgG B-cell clones were analyzed (**Figure 3C, left panels**). Reactivity patterns of IgG B-cell clones to SARS-CoV-2 S were similar to those detected before XBB.1.5 vaccination for all individuals. However, an increased proportion of IgG B-cell clones broadly reacted to all variant S (clones 1-157 [70%]) (**Figure 3D and Supplementary Figure 4C**). At 3-6 months after XBB.1.5 vaccination, a total of 133 S-reactive IgG B-cell clones were detected (**Figure 3C, left panels**). The proportion of clones that recognized all variants decreased slightly at 3-6 months compared to day 28 (84/133 [63%]; 1.11-fold reduction) (**Figure 3D**).

At baseline, 72/311 S-reactive IgG B-cell clones (23%) specifically targeted the RBD (**Figure 3C, right panels**). Broadly reactive IgG B-cell clones targeting RBD were also detected at that timepoint (clones 1-17 [24% of RBD-reactive clones]), albeit proportionally less than for the full S (**Figure 3D**). Instead, a large fraction of RBD-targeting clones cross-recognized ancestral, Delta and some but not all Omicron subvariants. Similar as for S, the number of broadly reactive IgG B-cell clones to RBD increased substantially at 28 days post-vaccination (clones 1-36 [55%]). At 3-6 months after booster, 52/133 S-reactive IgG B-cell clones (39%) targeted RBD and displayed similar reactivity profiles to those detected on day 28 post-vaccination (**Figure 3C, right panels**). However, the number of RBD-reactive clones that were broadly reactive to all variants decreased to a greater extent compared to S (22/52 [42%]; 1.31-fold reduction) (**Figure 3D**). A sporadic clone with unique XBB.1.5 RBD-specificity was detected 3-6 months after XBB.1.5 vaccination, but no other XBB.1.5 type-specific clones were detected in prior timepoints (**Figure 3C and Supplementary Figure 4D**). Nevertheless, broadly reactive IgG B-cell clones with binding capacity to all variant S, but also RBD, up to JN.1 were detectable in all individuals (**Figure 3C**). Additionally, we detected a gradual increase in the RBD-reactive clones that selectively recognized Omicron variants (**Figure 3D**).

### XBB.1.5 vaccination boosts Omicron-specific memory B-cells

Spectral flow cytometry was conducted on PBMC from 14 individuals to evaluate the memory B-cell (Bmem) response after monovalent XBB.1.5 vaccination. The overall Bmem compartment was similar pre– and 28 days post-XBB.1.5 vaccination (**Supplementary Figure 5 and 6**). Within Bmem, double-discrimination was used to identify ancestral SARS-CoV-2 (ANC) and XBB.1.5 RBD-binding or cross-reactive Bmem (**Figure 4A**). Three populations were defined: ANC^+^XBB.1.5^-^, ANC^+^XBB.1.5^+^, and ANC^-^XBB.1.5^+^ RBD-binding Bmem, with the latter making up the majority of RBD-reactive Bmem. Of note, these populations may cross-react with other variants. The proportions of ANC^+^XBB.1.5^-^ RBD-binding Bmem remained unchanged pre– and post-XBB.1.5 vaccination, while proportions of B-cells that were either ANC^+^XBB.1.5^+^ *(P =* 0.0006) or ANC^-^XBB.1.5^+^ (*P =* 0.0001) significantly increased from pre-to 28 days post-monovalent XBB.1.5 vaccination (**Figure 4A**).

**Figure 4.**
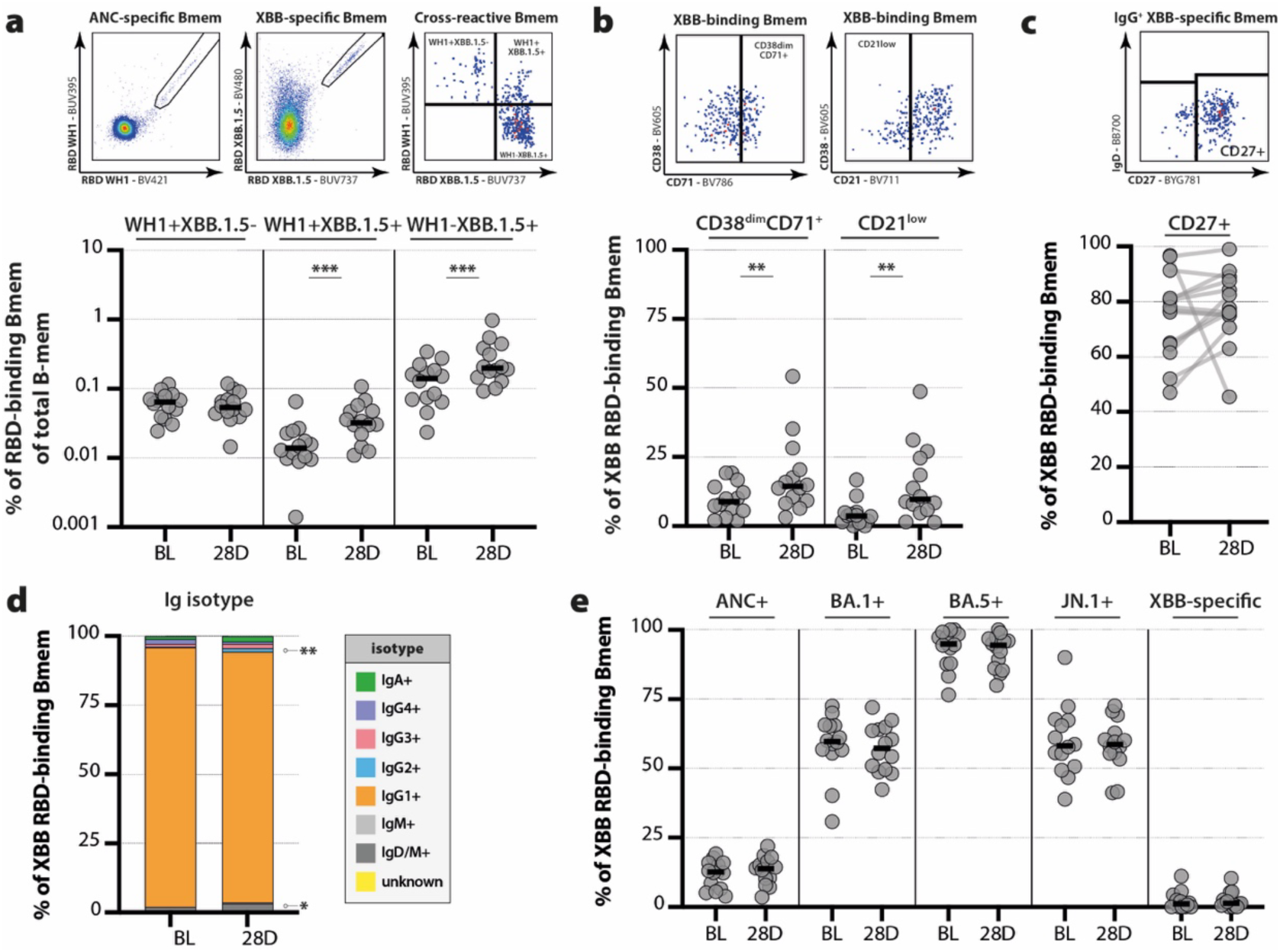
Cross-reactivity of RBD-specific memory B-cells. **(A)** Proportions of ancestral (ANC)+XBB.1.5-, ANC+XBB.1.5+ and ANC-XBB.1.5+ RBD-specific B-memory (B-mem) cells pre– and 28 days post-monovalent XBB.1.5 vaccination. Top panel represents the gating strategy used to identify ANC-specific B-mem (left), XBB.1.5-specific B-mem (middle), and cross-reactive B-mem (right) between the ancestral SARS-CoV-2 and XBB.1.5 variant. **(B)** CD38dimCD71+ and CD21lo cells within total XBB.1.5 RBD-specific Bmem. Top panel represents the gating strategy used to identify CD38dimCD71+ and CD21lo cells within the XBB.1.5 RBD-specific B-mem subset **(C)** CD27+ events within IgG+ XBB.1.5 RBD-specific Bmem. Top panel represents the gating strategy used to identify CD27+ cells within the IgG+ XBB.1.5 RBD-specific B-mem subset. **(D)** Distribution of Ig isotype and IgG subclass expressing subsets within XBB.1.5 RBD-specific Bmem. **(E)** Proportions of ancestral SARS-CoV-2, BA.1, BA.5, and JN.1 RBD-cross-reactive cells within XBB.1.5 RBD-specific Bmem. Non-cross-reactive cells determined to bind only XBB.1.5 RBD are shown on the right side of the graph. Statistical significance as determined by Wilcoxon matched-pairs signed rank test for paired data; \*\**P*<0.01, \*\*\**P*<0.001.

The activation profiles of all XBB.1.5 RBD-binding CD38^dim^ Bmem were investigated by determining expression levels of CD71 and CD21: upregulation of CD71 is indicative of recent activation, whilst a reduction in CD21 expression occurs upon antigen recognition. A significantly higher proportion of XBB.1.5 RBD-binding Bmem were CD71^+^ (*P =* 0.004) and CD21^lo^ (*P =* 0.0052) 28 days after XBB.1.5 vaccination (**Figure 4B**). The proportions of CD27^+^ cells within IgG^+^ XBB.1.5 RBD-binding Bmem were assessed further to measure secondary germinal center (GC)-experienced class-switched Bmem. There was a trend (*P* = 0.08) toward increased frequencies of CD27^+^IgG^+^ XBB.1.5 RBD-binding Bmem from pre-to 28 days post-monovalent XBB.1.5 vaccination (**Figure 4C**). Next, the Ig-isotype and IgG subclass distribution of XBB.1.5 RBD-binding Bmem were determined pre– and 28 days post-monovalent XBB.1.5 vaccination (gating in **Supplementary Figure 7A**). The vast majority of XBB.1.5 RBD-binding Bmem expressed IgG1 at both timepoints. However, there was a significant increase in the frequencies of IgM-only (*P =* 0.02) and IgG2 (*P =* 0.01) expressing XBB.1.5 RBD-binding Bmem from pre-to post XBB.1.5 vaccination (**Figure 4D**). There was also a trend (*P* = 0.08) toward reduced frequencies of IgG4 expressing XBB.1.5 RBD-binding Bmem from pre-to post XBB.1.5 vaccination.

Finally, the capacity of XBB.1.5 RBD-binding Bmem to cross-bind ancestral SARS-CoV-2 and other Omicron subvariants BA.1, BA.5, and JN.1 was evaluated (gating in **Supplementary Figure 7B**). The frequencies of XBB.1.5 RBD-binding Bmem that recognized other variants were similar between pre– and 28 days post-monovalent XBB.1.5 vaccination timepoints (**Figure 4E**). XBB.1.5 RBD-binding cells were found to have high cross-reactivity (90%) to BA.5 at both timepoints, suggesting these are formed after previous bivalent vaccination and/or omicron breakthrough infection. The frequencies of RBD-specific Bmem that only recognized the XBB.1.5 variant (and not other omicron variants) were low, in line with the clonal reactivity analysis (**Supplementary Figure 4D**). These frequences were also unchanged 28 days after XBB.1.5 vaccination. The effect of differences between previous bivalent booster vaccinations was evaluated by comparing variant-binding capacity between subjects who had received a BA.1 or BA.5 bivalent vaccine 12 month prior to XBB.1.5 vaccination. The frequencies of XBB.1.5 RBD-binding Bmem that could also bind BA.5 was higher in subjects that had previously received a BA.5 bivalent vaccine, both pre (*P =* 0.11) and 28 days (*P =* 0.04) post-XBB.1.5 vaccination (**Figure 4E**).

### Robust memory T-cell responses to emerging Omicron variants

The kinetics of ancestral SARS-CoV-2-specific T-cell responses were measured using an in-house developed IFNγ release assay (**Figure 5A**). Minimal increases were observed following monovalent XBB.1.5 vaccination, with geometric mean IFNγ levels increasing from 1.32 IU/mL at baseline to 1.80 IU/mL at day 28, before gradually declining back to baseline by 3-6 months. When evaluating response profiles with peptide pools covering S variants at 3-6 months after monovalent XBB.1.5 vaccination, T-cell responses showed robust cross-reactivity (**Figure 5A**). However, in about 20% of individuals the T-cell response to circulating Omicron variants (BA.1, BA.2, BA.5, BQ.1.1, XBB.1.5 and EG.5.1) was reduced to approximately 80% reactivity compared to ancestral (**Supplementary Figure 8**). T-cell responses to common cold coronaviruses (CCC; NL63 and OC43) were readily detected albeit at a lower level than SARS-CoV-2-specific responses, aligning with recent multiple exposures to SARS-CoV-2 S as compared to CCC S. MERS-CoV-specific responses were detected at low level in approximately 30% of participants. Since participants were not exposed to MERS-CoV, these are likely cross-reactive T-cell responses.

**Figure 5.**
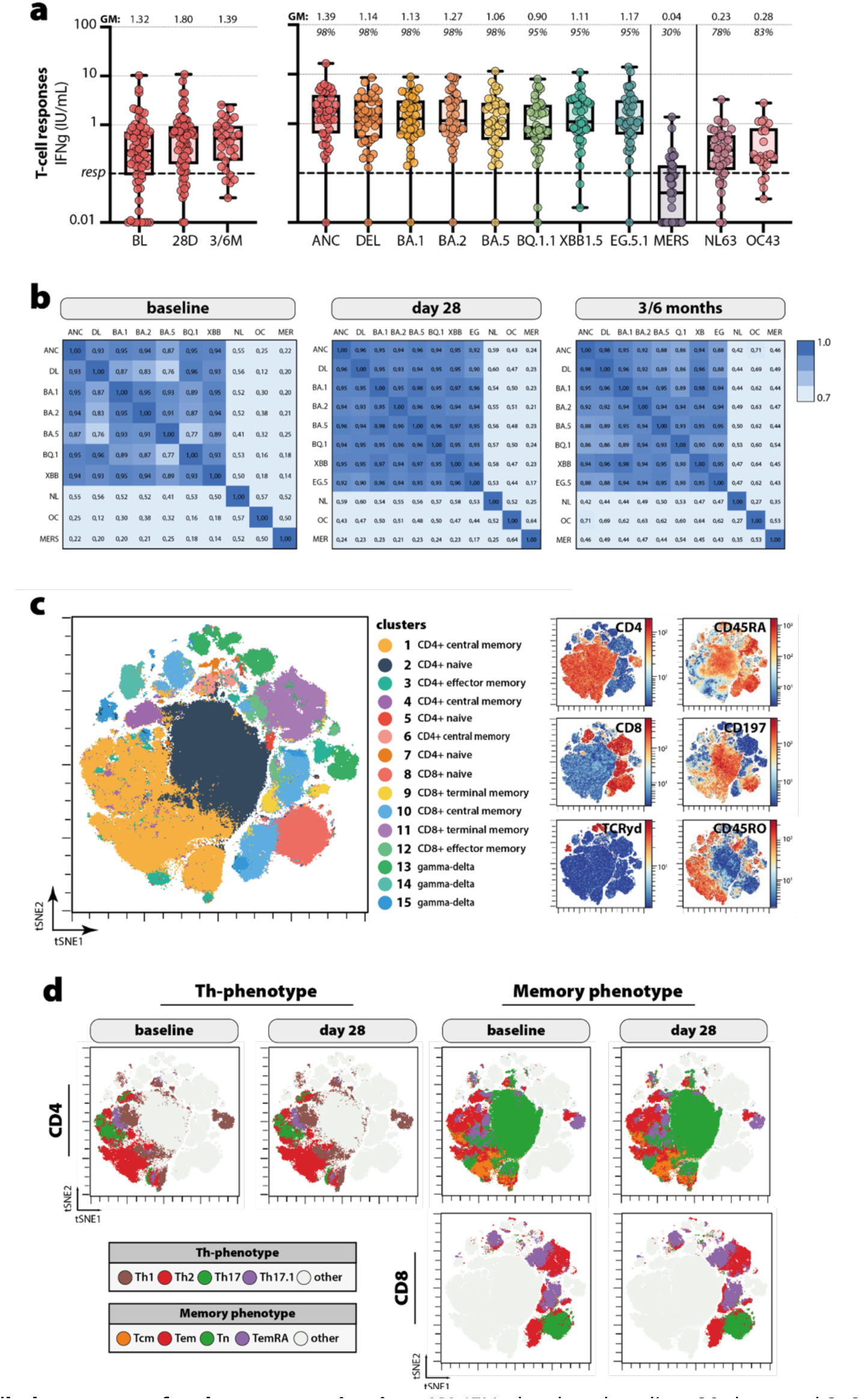
T-cell phenotypes after booster vaccination. **(A)** IFNg levels at baseline, 28 days, and 3-6 months post XBB.1.5 vaccination after stimulation of whole blood with a peptide pool covering ancestral SARS-CoV-2 (left), and T-cell profiles at 3-6 months after XBB.1.5 vaccination for ancestral, Delta, BA.1, BA.2, BA.5, BQ.1.1, XBB.1.5, EG.5.1, MERS-CoV, NL63, and OC43 (right). **(B)** Spearman correlation matrix between Delta (DL), BA.1 (B.1), BA.2 (B.2), BA.5 (B.5), BQ.1.1 (Q.1), XBB.1.5 (XB), ancestral (ANC) SARS-CoV-2, and CCC NL63 (NL), OC43 (OC) and MERS-CoV (MER) at baseline-, 28-, and 3-6-months post XBB.1.5 vaccination. **(C and D)** FlowSOM cluster analysis (optSNE) and clustered heatmap of LIVE CD3+ T-cells in a multicolor CyTOF T-phenotyping panel. The horizontal lines of the box-and-whisker plots indicate the median, the whiskers indicate the range, and the bounds of the boxes indicate the interquartile range (IQR). All individual datapoints are shown for the individuals in the box-and-whisker plots.

To better assess the breadth of the SARS-CoV-2-specific T-cell response over time, median IFNγ responses between SARS-CoV-2 variants, CCC, and MERS-CoV were correlated at baseline, 28 days, and 3-6 months after monovalent XBB.1.5 booster vaccination. SARS-CoV-2 variant responses exhibited strong correlation at baseline. An increased correlation between SARS-CoV-2 variant IFNγ responses was seen 28 days after booster vaccination, which was retained at to 3-6 months (**Figure 5B**).

Immunophenotyping using a 38-marker CyTOF panel revealed that 28 days post-monovalent XBB.1.5 vaccination, overall immune cell frequencies (**Supplementary Figure 9A**) or T-cell phenotypes (**Supplementary Figure 9B**) were not altered. Further analysis of T-cells using unsupervised flowSOM clustering showed five main phenotypes: naïve CD4 T-cells (cluster 2, 5, and 7), memory CD4 T-cells (cluster 1, 3, 4, and 6), naïve CD8 T-cells (cluster 8), memory CD8 T-cells (cluster 9 to 12), and gamma-delta T-cells (cluster 13 to 15) based on differential marker expression (**Figure 5C and Supplementary Figure 10 and 11**). CD4 and CD8 T-cell clusters could be further divided into different clusters of T-helper phenotypes (CD4; Th1, Th2, Th17, and Th17.1, **Figure 5D and Supplementary Figure 10 and 11**) and memory phenotypes (CD4 and CD8; T-naive [Tn], T-central memory [Tcm], T-effector memory [Tem], and T-terminal memory [TemRA]). CD4+ T-helper phenotypes (i.e., Th1, Th2, Th17, and Th17.1) and CD4+ and CD8+ memory phenotypes were manually gated and overlayed on the optSNE and confirmed the phenotype of FlowSOM clusters (**Figure 5D**).

AIM+CD4+ and AIM+CD8+ (CD137+CD69+) T-cells were manually gated and overlayed on the optSNE. Most SARS-CoV-2-specific memory CD4+ T-cells exhibited a Th2-like (cluster 4; CXCR3-CCR4+CCR6-) phenotype, while CD8+ T-cells mostly presented a terminal memory (cluster 11; CD45RA-, CD197-) phenotype (**Figure 6A**). Interestingly, a large proportion of S-specific CD8⁺ memory T-cells clustered in clusters 11 and 12, which were characterized by high CD57 expression (**Supplementary Figure 10**), indicating a senescent phenotype of SARS-CoV-2-specific CD8⁺ T-cells. However, no significant changes in the phenotype or frequencies were detected 28 days after vaccination. Additionally, ancestral and XBB.1.5-specific T-cell responses demonstrated similar phenotypes and were equally present in peripheral blood. In contrast to the IGRA, no increases in SARS-CoV-2-specific T-cess were detected post vaccination, (**Figure 6B**, **Supplementary Figure 12** for individual participants).

**Figure 6.**
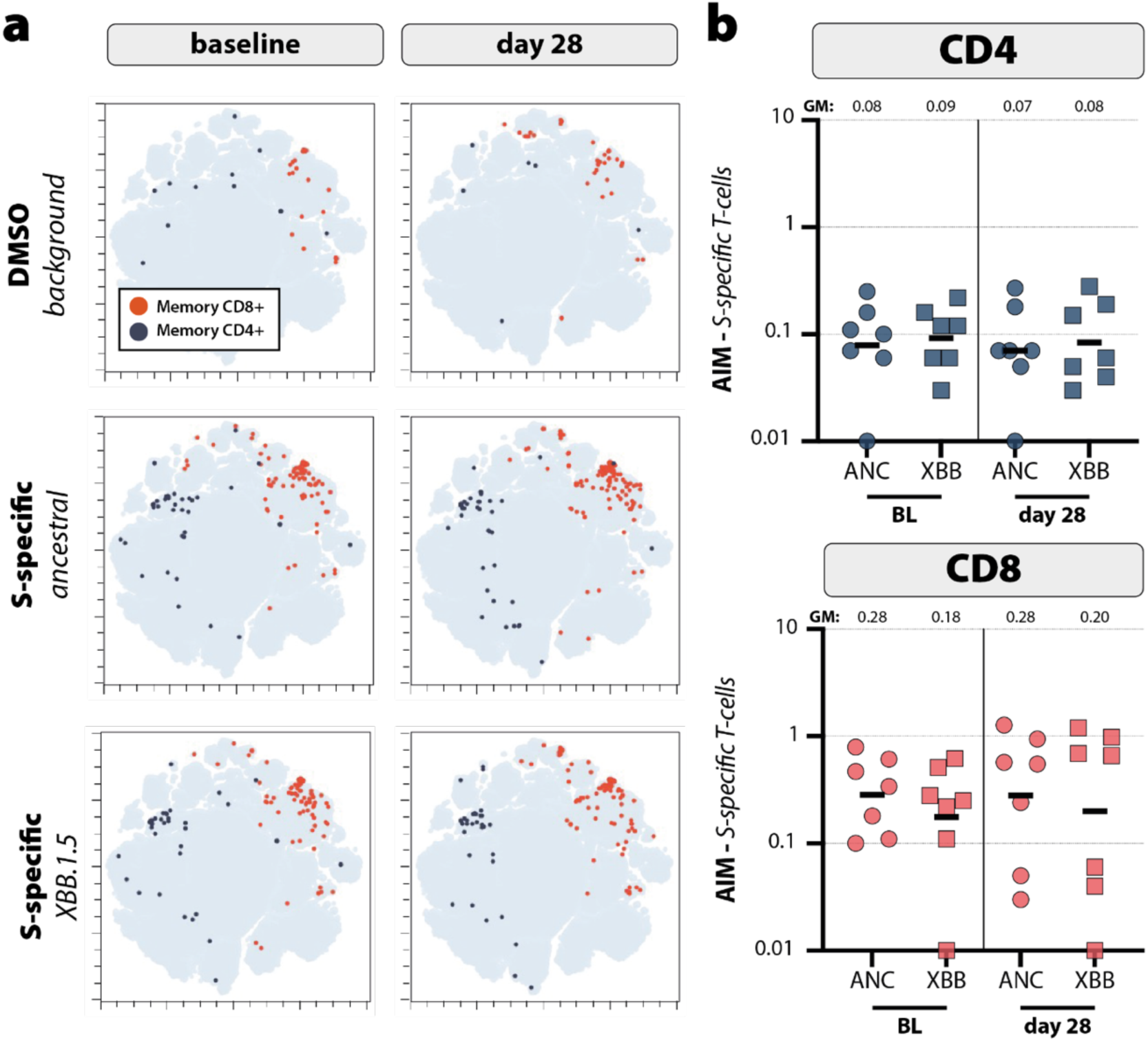
S-specific T-cell responses after booster vaccination. **(A)** Overlay of CD4+ (blue) and CD8+ (red) AIM+ (CD137+CD69+) T-cells on the FlowSOM cluster analysis (optSNE) at baseline and 28 post XBB.1.5 vaccination. **(B)** AIM+ CD4+ and CD8+ T-cells at baseline and 28 for the ancestral and XBB.1.5 S using manual gating. Geometic mean IFNg levels or AIM frequencies are shown on top of each box. The horizontal line represents the geometric mean. All individual datapoints are shown.

## Discussion

In this study, we performed in-depth immune profiling to assess the magnitude, breadth, and phenotype of virus-specific immune responses following monovalent XBB.1.5 booster vaccination in healthcare workers. Monovalent XBB.1.5 vaccination boosted binding, neutralizing, and ADCC-mediating antibody responses with broad-reactivity to emerging Omicron subvariants. Although antibody titers against XBB.1.5 and other Omicron subvariants increased preferentially, responses remained dominated by recognition of the ancestral S. Clonal analysis of CD19+ IgG B-cells confirmed that these antibody responses were primarily mediated by the recall of broadly reactive B-cell clones, with minor induction of clones selectively recognizing Omicron variants. This explains the persistent immunodominance of early SARS-CoV-2 strains and broad boost of antibody titers observed, demonstrating the presence of immune imprinting. Regardless, within the Bmem subset XBB.1.5 RBD-binding cells were preferentially boosted. T-cells were broadly cross-reactive and showed minimal changes in magnitude and phenotype post-vaccination. Overall, the monovalent XBB.1.5 booster effectively reinforced pre-existing immune memory and skewed this towards increased recognition to the vaccine variant.

Neutralizing antibodies are considered the main correlate of protection against (severe) COVID-19, and a potential immunological readout of vaccine efficacy (*27, 28*). Low neutralizing antibody levels increase the risk of breakthrough infections with antigenically distinct variants (*29*). While Omicron-specific binding and functional antibody levels were lower than those directed to the ancestral strain in this study, neutralization titers to emerging Omicron subvariants were boosted by XBB.1.5 vaccination. Not only did XBB.1.5 booster vaccination lead to a 5-fold increase in XBB.1.5 neutralization, it also increased neutralization of variants that emerged later, like BA.2.86 and EG.5.1, proving that monovalent vaccination can induce reactivity of neutralizing antibodies beyond the vaccine strain and towards novel antigenically similar variants (*30*).

Mutations in the S protein that lead to evasion from neutralizing antibodies had less effect on the activity of ADCC-mediating antibodies. This is because antibodies with Fc-effector functions also bind more conserved epitopes in S outside of the RBD (*31*), allowing for broader recognition of emerging SARS-CoV-2 variants that harbor RBD-associated immune evasive mutations (*32, 33*). Monovalent XBB.1.5 vaccination increased ADCC-mediating antibody levels about 2-to 3-fold, and XBB.1.5-specific titers were similar to those directed to the Omicron BA.1 and BA.5 subvariants, and only about 2-fold lower than those targeting ancestral S.

Although we did not detect significant differences in the immunogenicity of XBB.1.5 monovalent vaccination in individuals with different prime-boost histories, binding and neutralizing antibody levels were in general higher in mRNA-primed than in Ad26.COV2.S-primed participants; consistent with previous reports (*34, 35*). In contrast, ADCC-mediating antibody levels were higher in Ad26.COV2.S-primed individuals. This indicates that the original priming history of the study participants still affects total antibody levels but also has distinct effects on antibody functionality. This could potentially be explained by IgG subclasses: repeated mRNA-based vaccination was associated with an increase in serum IgG4 isotypes and IgG4 memory B-cells, explaining the lower fraction of antibodies with Fc-mediated functionality (*36*). However, we observed only a minor fraction of circulating XBB.1.5-binding IgG4 Bmem compared to IgG1, but we did not perform subclass analyses of serum binding IgG antibodies to confirm this finding. Of note, our in-depth RBD-binding Bmem phenotyping was mainly performed on samples from Ad26.COV2.S-primed individuals due to limited sample availability, potentially explaining this discrepancy.

Serum antibody levels to all tested variants were boosted by monovalent XBB.1.5 vaccination. Indeed, a minimal number of B-cell clones with unique reactivity to XBB.1.5 were detected and broadly reactive B-cells targeting S were primarily boosted by vaccination. When assessing reactivity with the less conserved RBD, we detected an increase in IgG B-cells that specifically targeted Omicron subvariants. Interestingly, at 3-6 months after vaccination, RBD-reactive clones waned to a lesser extent compared to the total S-reactive IgG B cell clones, and clones that reacted towards RBD of more recent omicron variants such as BA.5, BQ.1.1, XBB.1.5 and BA.2.86 waned least. Together, these data suggest that despite the dominance of ancestral imprinted broadly reactive IgG clones, a fraction of Omicron RBD-reactive clones is gradually selected over time. This shows that immune imprinting by previous exposures from infection and/or vaccination dominates the variant response (*37*), yet does not impair reshaping of immune memory towards variant recognition (*38, 39*). Our IgG B-cell reactivity analysis includes a wide range of IgG B-cell phenotypes but cannot discriminate circulating IgG clones that are formed *de novo* from those that are re-called from prior exposures. However, through phenotypic analysis of RBD-specific Bmem, we found that both XBB.1.5 RBD-reactive and cross-reactive ancestral-XBB.1.5 RBD Bmem displayed an activated phenotype and were preferentially boosted, and not those specifically recognizing the ancestral variant. Together, our data indicates that upon XBB.1.5 booster vaccination, a fraction of IgG B cells with reactivity to XBB.1.5 are preferentially boosted, counterbalancing prior imprinting.

While mutations in S of emerging Omicron variants result in partial immune escape from neutralizing antibody responses, T-cell responses retain variant-wide reactivity (*16, 41*). Monovalent XBB.1.5 vaccination only modestly boosted SARS-CoV-2-specific T-cell responses as measured by IGRA, with a slight increase observed 28 days-post vaccination. However, compared to CCC-specific T-cell responses, SARS-CoV-2-specific T-cell responses were already high at baseline, suggesting that these memory pools may be near saturation and therefore have limited capacity for further expansion upon booster vaccination, which is supported by a senescent phenotype of a proportion of reactive CD8+ T-cells. Booster vaccination did enhance the cross-reactivity profile of T-cell responses, broadening the T-cell reactivity landscape by day 28 and sustaining this increase up to 3-6 months after vaccination. However, a reduction in cross-recognition of Omicron variants by T-cells was observed in ±20% of individuals, which might be explained by specific HLA-profiles vulnerable for mutations in Omicron S (*42, 43*).

Immune phenotyping revealed no changes in overall immune cell frequencies or T-cell phenotypes post-vaccination, with SARS-CoV-2-specific CD4+ T-cells showing a dominant Th2-like profile and CD8+ T-cells having mainly a terminal memory phenotype. Interestingly, S-specific CD8+ T-cells often highly expressed CD57; this marker is found at the end of the differentiation pathway and related to repeated antigen exposure. Even though these cells have little proliferative capacity, they are thought to maintain their cytotoxic potential and are therefore regarded senescent rather than exhausted (*44*). This could explain the only modest increase in T-cell responses post monovalent XBB.1.5 booster vaccination observed in IGRA, which could not be confirmed by flow cytometry, indicating that there is limited capacity for further expansion post-vaccination. Since both ancestral and XBB.1.5-specific T-cell responses were comparably boosted and maintained similar phenotypes, this shows that booster vaccination reinforced T-cell immunity but does not distinctly favor induction of responses to a specific virus strain.

Even though some analyses could only be performed in a small subset of individuals, all data suggest that monovalent XBB.1.5 vaccination reinforces cross-reactive immune memory, particularly through recall of broadly reactive B-cells and preservation of robust T-cell responses. Omicron RBD-specific B-cell memory was preferentially boosted, with limited *de novo* induction of type-specific B-cell clones. This suggests imprinted memory responses shape responses to variant-targeted boosters, but do not restrict redirecting the memory B-cell response. However, antigenically distinct SARS-CoV-2 immune escape variants will continue to emerge and monitoring its impact in vaccination strategies, especially in risk groups, remains of crucial importance.

## Data availability statement

All data produced in the present study are available upon reasonable request to the authors.

## Acknowledgments

We thank the ARAFlowCore staff at Melbourne for staining and assistance with flow cytometry.

## Funding statement

The bivalent BA.5 vaccine mRNA-1273.222 was provided by Moderna. Moderna had no role in study design, data collection, data analysis, data interpretation, or writing of the report. All other vaccines were supplied by the Center for Infectious Disease Control, National Institute for Public Health and the Environment, the Netherlands (RIVM). This study was funded by the Netherlands Organization for Health Research and Development (ZonMw), grant agreements 10430072110001 (P.H.M.vdK., C.H.GvK., R.D.dV.) and 10430072110008 (C.H.GvK., R.D.dV.). The funder of the study had no role in study design, data collection, data analysis, data interpretation, or writing of the report.

Daryl Geers^1*^, Muriel Aguilar-Bretones^1*^, Luca M. Zaeck^1*^, Paul A. Gill^2$^, Cristina Gonzalez-Lopez^1$^, Kim Handrejk^1$^, Ngoc H. Tan^3$^, Yvette den Hartog^4^, Lennert Gommers^1^, Susanne Bogers^1^, Anna Z. Mykytyn^1^, Roos S.G. Sablerolles^3^, Abraham Goorhuis^5,6^, Douwe F. Postma^7^, Leo G. Visser^8^, Virgil A.S.H. Dalm^9^, Melvin Lafeber^10^, Neeltje A. Kootstra^11^, Debbie van Baarle^12,13^, Bart L. Haagmans^1^, Marion P.G. Koopmans^1^, Corine H. GeurtsvanKessel^1#^, P. Hugo M. van der Kuy^3#^, Gijsbert P. van Nierop^1#^, Menno C. van Zelm^2,14#^, Rory D. de Vries^1§^

