## Supplemental figures for "Preferential boosting of SARS-CoV-2 Omicron lineage-specific immune responses by monovalent XBB.1.5 vaccination"

[1] Department of Viroscience, Erasmus MC, Erasmus University Medical Center, Rotterdam, the Netherlands
[2] Department of Immunology, School of Translational Medicine, Monash University, Melbourne, VIC, Australia.
[3] Department of Hospital Pharmacy, Erasmus MC, Erasmus University Medical Center, Rotterdam, the Netherlands
[4] Department of Transplantation, Erasmus MC, Erasmus University Medical Center, Rotterdam, the Netherlands
[5] Center of Tropical Medicine and Travel Medicine, Department of Infectious Diseases, Amsterdam University
[6] Medical Centers, Amsterdam, the Netherlands Infection & Immunity, Amsterdam Public Health, University of Amsterdam, Amsterdam, the Netherlands
[7] Department of Internal Medicine and Infectious Diseases, University Medical Center Groningen, Groningen, the Netherlands
[8] Department of Infectious Diseases, Leiden University Medical Center, Leiden, the Netherlands
[9] Department of Internal Medicine, Division of Allergy & Clinical Immunology and Department of Immunology, Erasmus Medical Center, Rotterdam, the Netherlands
[10] Department of Internal Medicine, Erasmus MC, Erasmus University Medical Center, Rotterdam, the Netherlands
[11] Department of Experimental Immunology, Amsterdam University Medical Centers, Amsterdam Infection and Immunity Institute, University of Amsterdam, Amsterdam, the Netherlands
[12] Department of Medical Microbiology and Infection Prevention, University Medical Center Groningen, University of Groningen, Groningen, the Netherlands
[13] Center for Infectious Disease Control, National Institute for Public Health and the Environment, Bilthoven, the Netherlands
[14] Department of Immunology, Erasmus MC, Erasmus University Medical Center, Rotterdam, the Netherlands
*,$,# These authors contributed equally

**12 Supplementary Figures**
**3 Supplementary Tables**

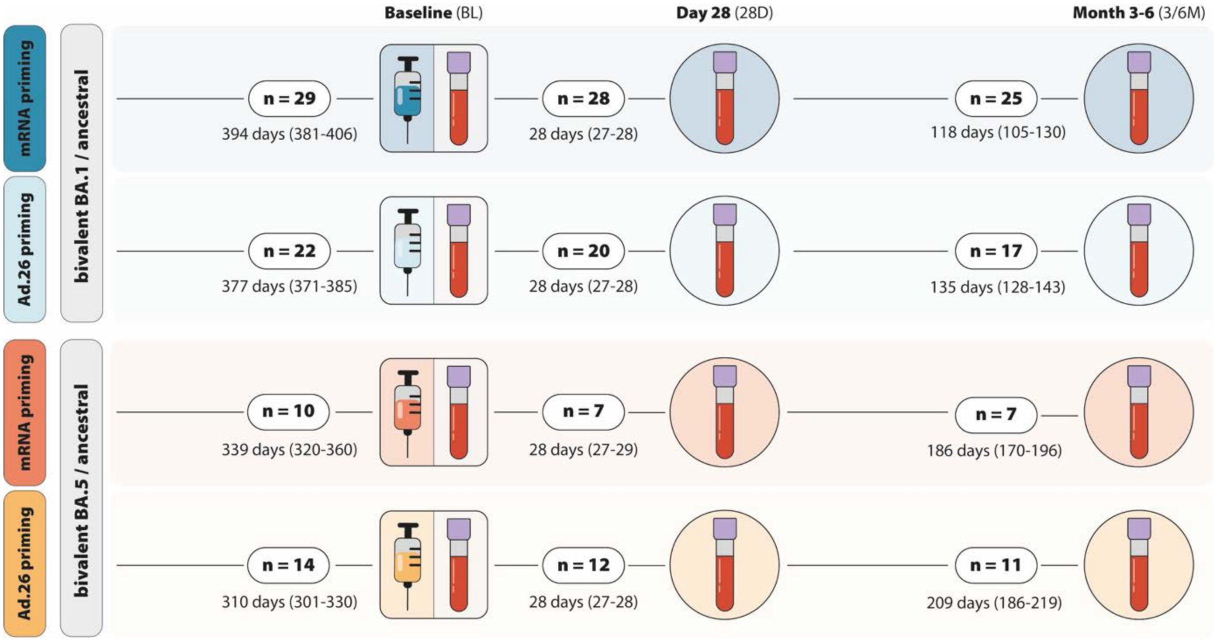


**Supplementary Figure 1. Study design.** Participants were enrolled from the SWITCH-ON study, have been primed with either mRNA-based vaccines or Ad26.COV2.S, and boosted with BA.1 bivalent or BA.5 bivalent vaccines. Twelve months following bivalent BA.1 or BA.5 vaccination, a fraction of participants were vaccinated with the monovalent XBB.1.5. At baseline (BL), 28 days (28D), and 3-6 months (3/6M) after monovalent XBB.1.5 vaccination, blood samples were collected for immunogenicity testing. Per group, the number of participants and time between sampling with interquartile range is indicated.

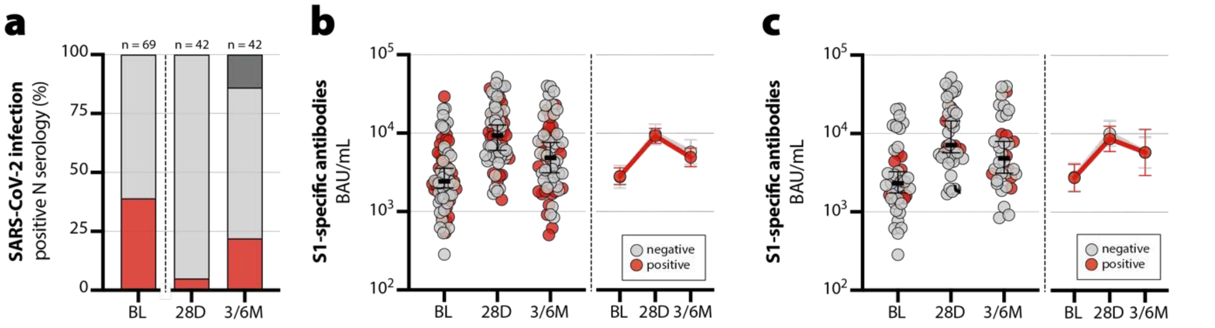


**Supplementary Figure 2. Breakthrough infections during the booster study.** Breakthrough infections were identified through N serology at BL, 28D, and 3/6M after monovalent XBB.1.5 vaccination. **(A)** Percentage of N positive participants (>1.4 S/CO ratio) are indicated for each timepoint; N-positive (red), N-negative (light gray), not tested (dark gray). **(B and C)** S1-specific IgG levels are shown for participants with or without breakthrough infection **(B)** before or **(C**) after XBB.1.5 vaccination (N- (positive, (red) in comparison to N-negative individuals. Individual datapoints are shown for all individuals, with the horizontal lines representing the geometric mean titers (GMT) and the whiskers indicating the 95% confidence intervals (CI). Line graphs next to each panel depict the GMT with 95% CI per group (N-positive [red] and N-negative [gray]).

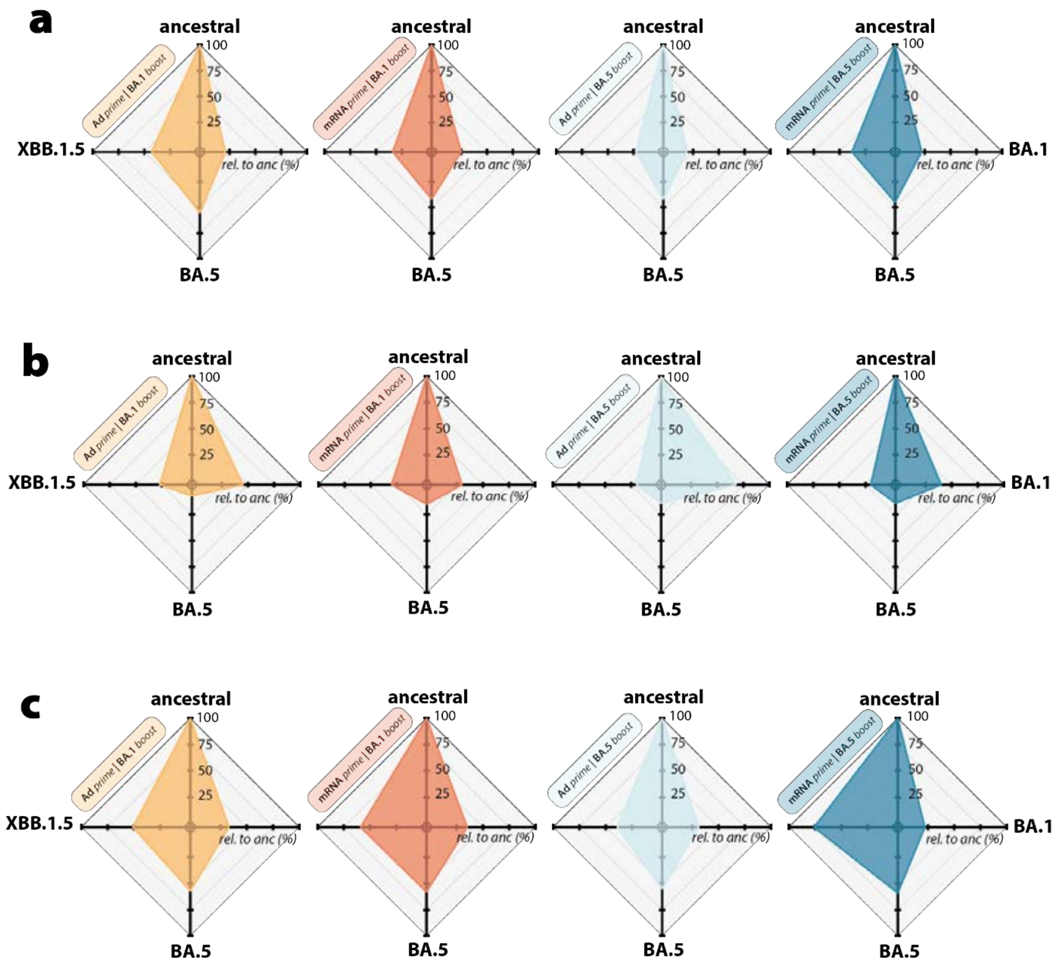


**Supplementary Figure 3. Relative functional antibody levels for Omicron variants.** Radar plots depicting variant-specific **(A)** binding, **(B)** neutralizing, and **(C)** ADCC-mediating antibody levels relative to ancestral SARS-CoV-2 per prime-boost regimen at 28 days post-vaccination.

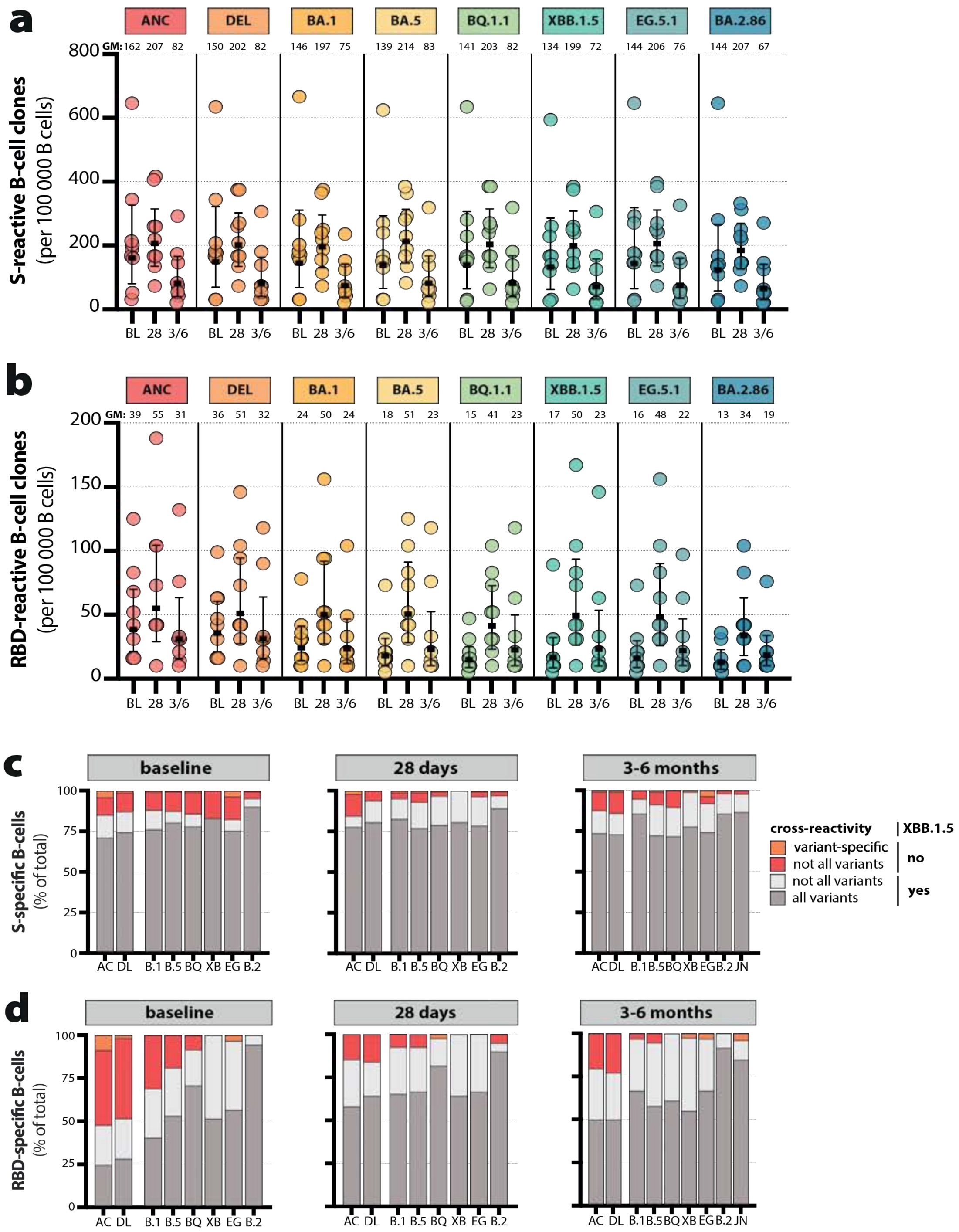


**Supplementary Figure 4. Individual S and RBD-specific B-cell frequencies and cross-reactivity fractions per variant. (A)** Individual SARS-CoV-2-reactive B-cell frequencies per 100,000 PMA-screened B-cells targeting the S trimer and **(B)** RBD at baseline (BL), 28 days (28D) and 3/6 months (3/6M) post-XBB.1.5 booster, color coded per variant **(C)** Fraction of IgG clones per timepoint that cross-recognize all variant S trimers (dark gray), that do not cross-recognize all variants but can cross-recognize XBB.1.5 (light grey), that do not cross-recognize XBB.1.5 (red) or that are variant specific (orange). **(D)** Fraction of RBD-specific IgG classified for broad variant reactivity (dark gray), cross-reactive to XBB.1.5 (light gray), not cross-reactive to XBB.1.5 (red) or variant specific (orange).

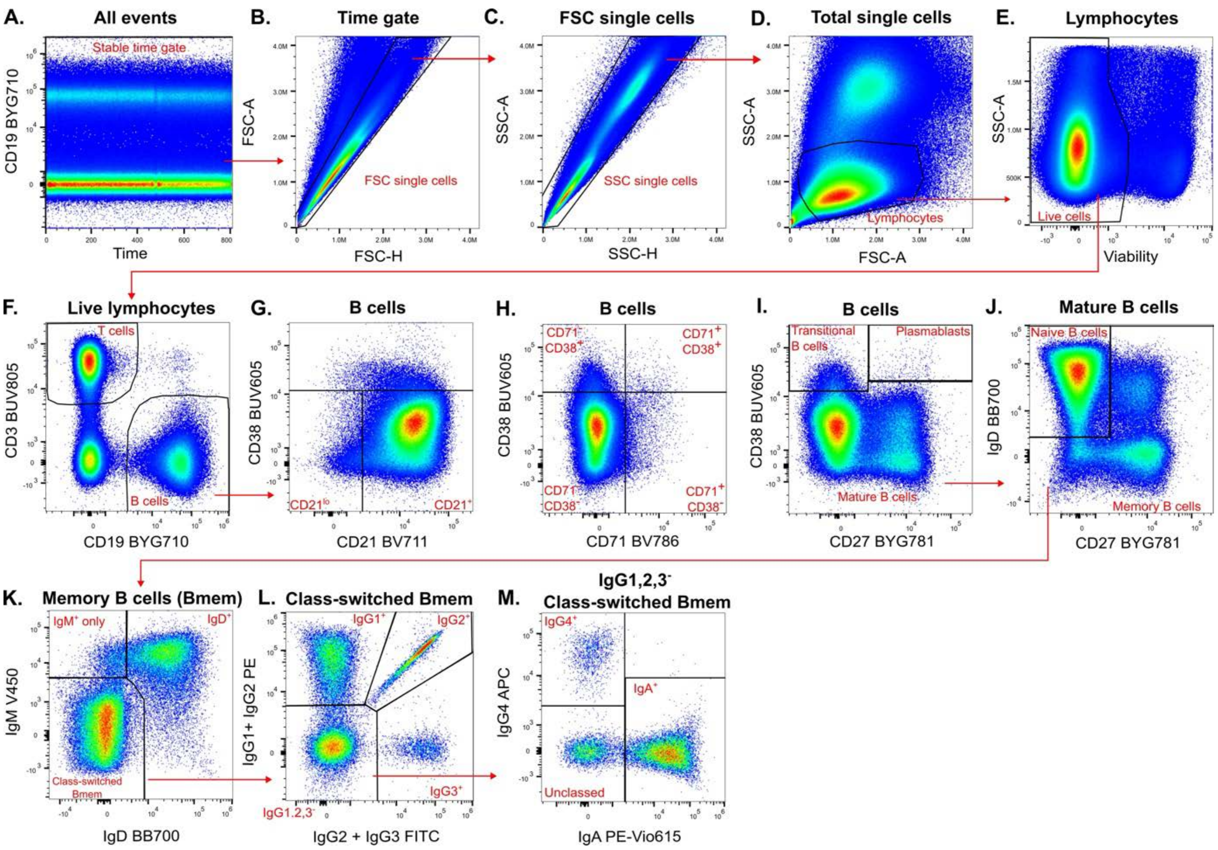


**Supplementary Figure 5. Memory B-cell (Bmem) gating strategy. (A)** All events were gated on CD19 vs time to ensure a steady signal across acquisition time and to exclude aggregates. **(B-C)** Doublets were excluded on FSC-A vs FSC-H, and subsequently SSC-A vs SSC-H. **(D)** Lymphocytes were gated as $SSC^{lo}FSC^{mid}$. **(E)** Dead cells were excluded on SSC-A vs viability. **(F)** B-cells were gated as $CD19^{+}CD3^{-}$. **(G)** $CD21^{lo}$ and $CD21^{+}$ B-cells were gated vs CD38. **(H)** CD38 and CD71 expression on B-cells was gated in quadrants. **(I)** Mature and transitional B-cells and plasmablasts were gated on CD38 vs CD27. **(J)** Bmem and naive B-cells were gated using IgD vs CD27 within mature B-cells. Bmem are additionally phenotyped on CD38, CD21 and CD71 expression using the same gating strategy shown in panels G and H. **(K)** Ig class-switched, $IgM^{+}IgD^{+}$, and $IgM^{+}$ only Bmem were gated on IgM vs IgD. **(L)** $IgG1^{+}$, $IgG2^{+}$, $IgG3^{+}$, and $IgG1,2,3^{-}$ class-switched mature Bmem were gated. **(M)** $IgG4^{+}$, $IgA^{+}$, and unclassed mature Bmem were gated within $IgG1,2,3^{-}$ cells.

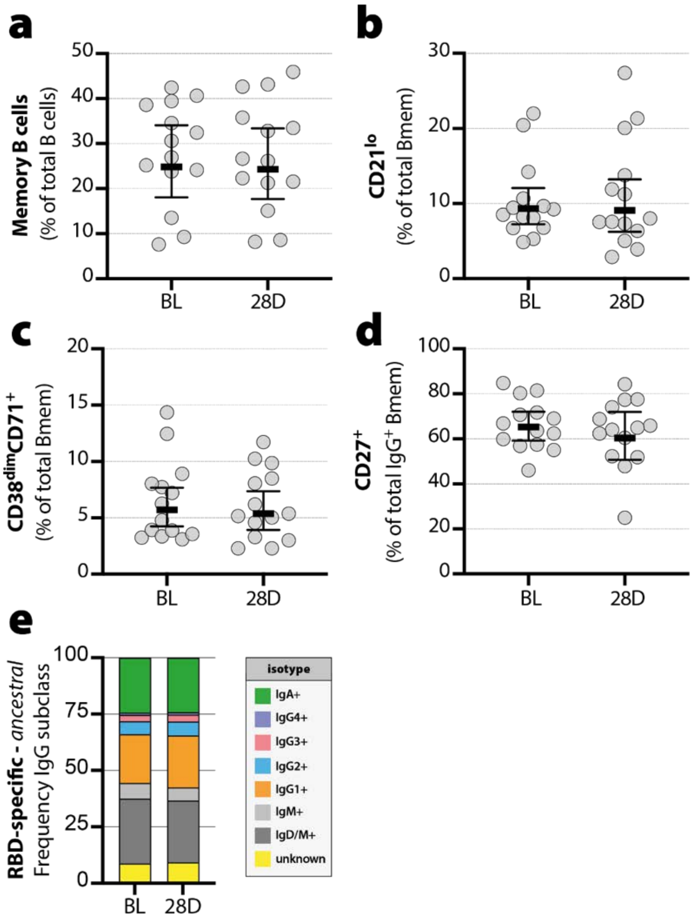


**Supplementary Figure 6. Memory B-cell (Bmem) phenotyping pre- and post-XBB.1.5 monovalent vaccination. (A)** Bmem as a proportion of total B-cells pre- and 1 month-post XBB.1.5 monovalent vaccination. **(B)** The proportion of recently activated CD21$^{lo}$ Bmem cells expressed as a percentage of total Bmem. **(C)** The proportion of recently activated CD38$^{dim}$CD71$^{+}$ Bmem cells expressed as a percentage of total Bmem. **(D)** The proportion of secondary germinal center experienced CD27+ Bmem expressed as a percentage of IgG$^{+}$ Bmem. **(E)** Distribution of Ig isotype and IgG subclass expressing subsets within total Bmem. Horizontal lines indicate the geometric mean and the whiskers indicate the 95% confidence interval. No statistically significant differences were identified (Wilcoxon matched pairs signed rank test).

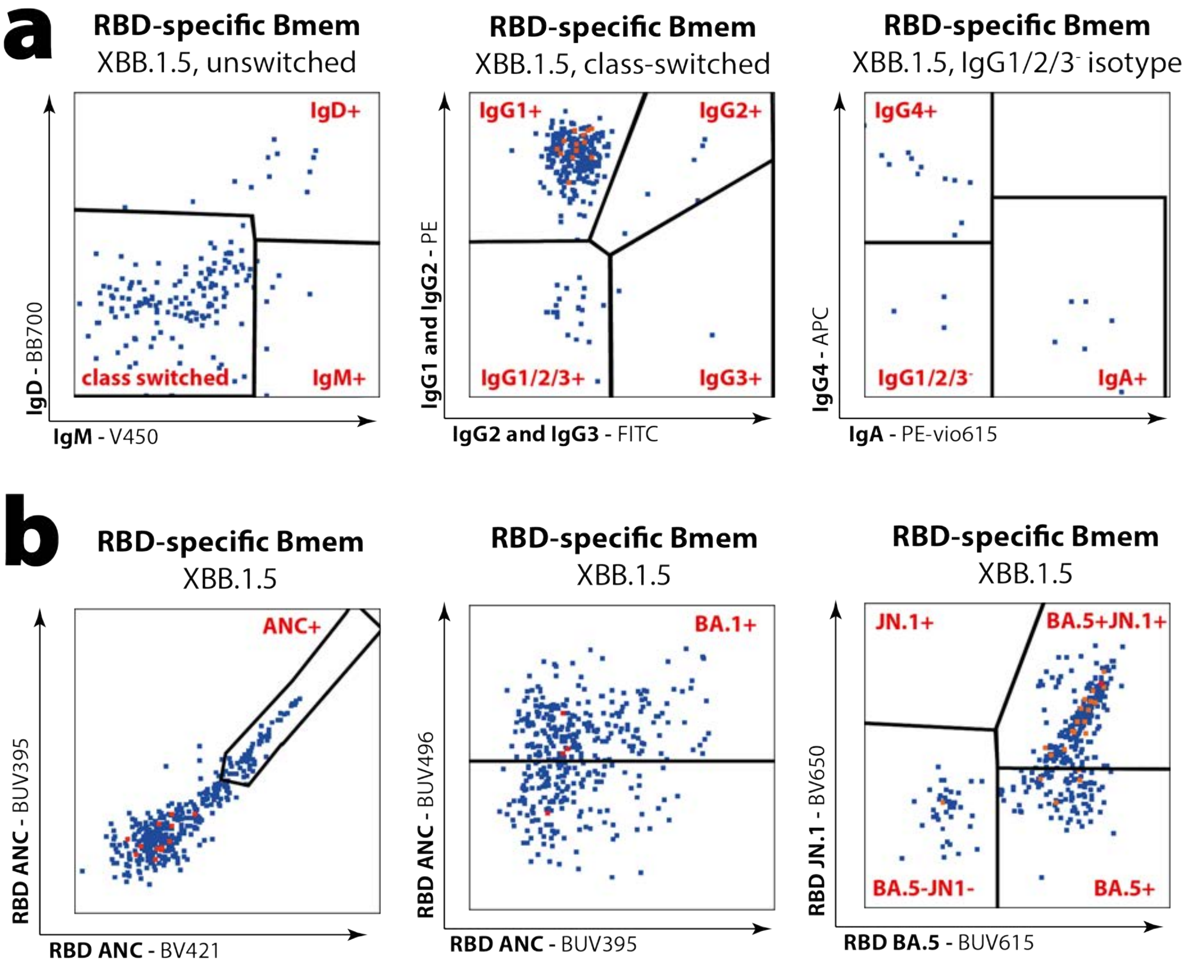


**Supplementary Figure 7. Gating strategy for Ig isotypes and subvariant binding within XBB.1.5 RBD-specific memory B-cells. (A)** XBB.1.5 RBD-specific memory B-cells (Bmem) were gated on IgD vs IgM expression to determine Ig class-switched, IgM+IgD+, and IgM+ only populations. Ig class-switched cells were then further subdivided into IgG1+, IgG2+, IgG3+ and IgG1,2,3-. Finally, IgG4+, IgA+ and unclassed XBB.1.5 RBD-specific Bmem were gated within IgG1,2,3- cells. **(B)** Gating strategy for detection of ANC, Omicron BA.1, BA.5 and JN.1 RBD within XBB.1.5 RBD-specific Bmem. Events that did not bind other variants were labelled XBB.1.5+ only.

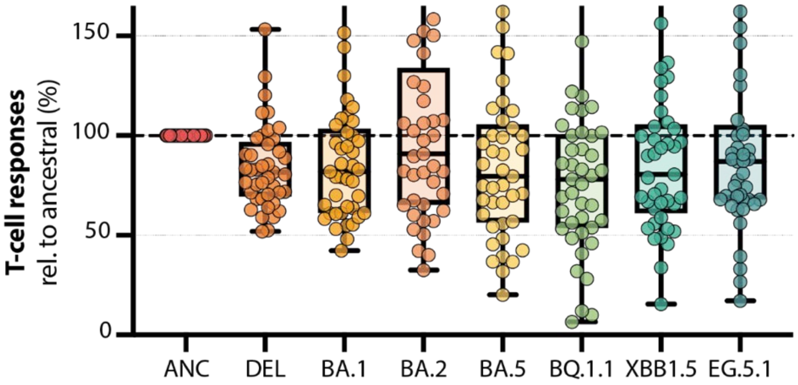


**Supplementary Figure 8. Relative T-cell responses to SARS-CoV-2 variants.** Percentage of IFNg levels relative to ancestral for the Delta, Omicron BA.1, BA.2, BA.5, BQ.1.1, XBB.1.5, and EG.5.1 variants. The horizontal dotted line indicates the 100% point. The horizontal lines of the box-and-whisker plots indicate the median, the whiskers indicate the range, and the bounds of the boxes indicate the interquartile range (IQR). All individual datapoints are shown for the individuals in the box-and-whisker plots.

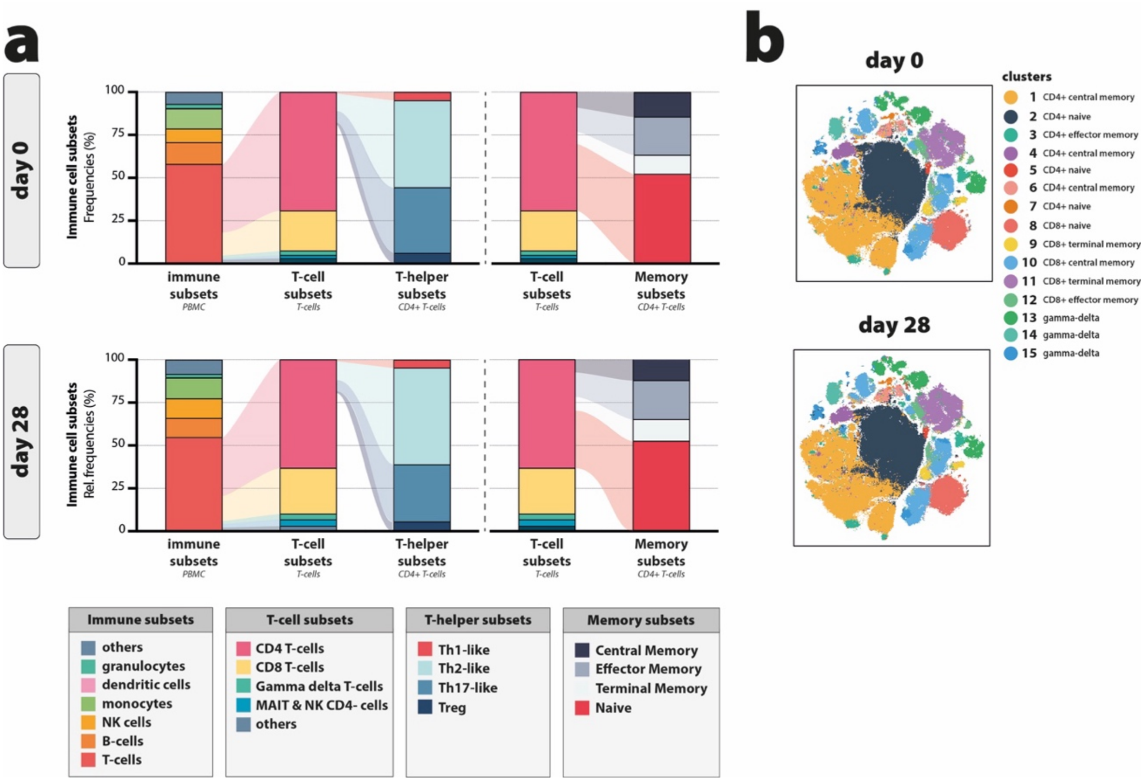


**Supplementary Figure 9. Phenotyping of PBMC pre and 28 days post vaccination. (A)** Relative frequencies of different immune subsets, T-cell subsets, T-helper subsets, and T-memory subsets at baseline and 28 days after XBB.1.5 vaccination for 9 paired individuals. **(B)** T cell phenotypes at baseline and day 28 after XBB.1.5 vaccination for the same paired individuals.

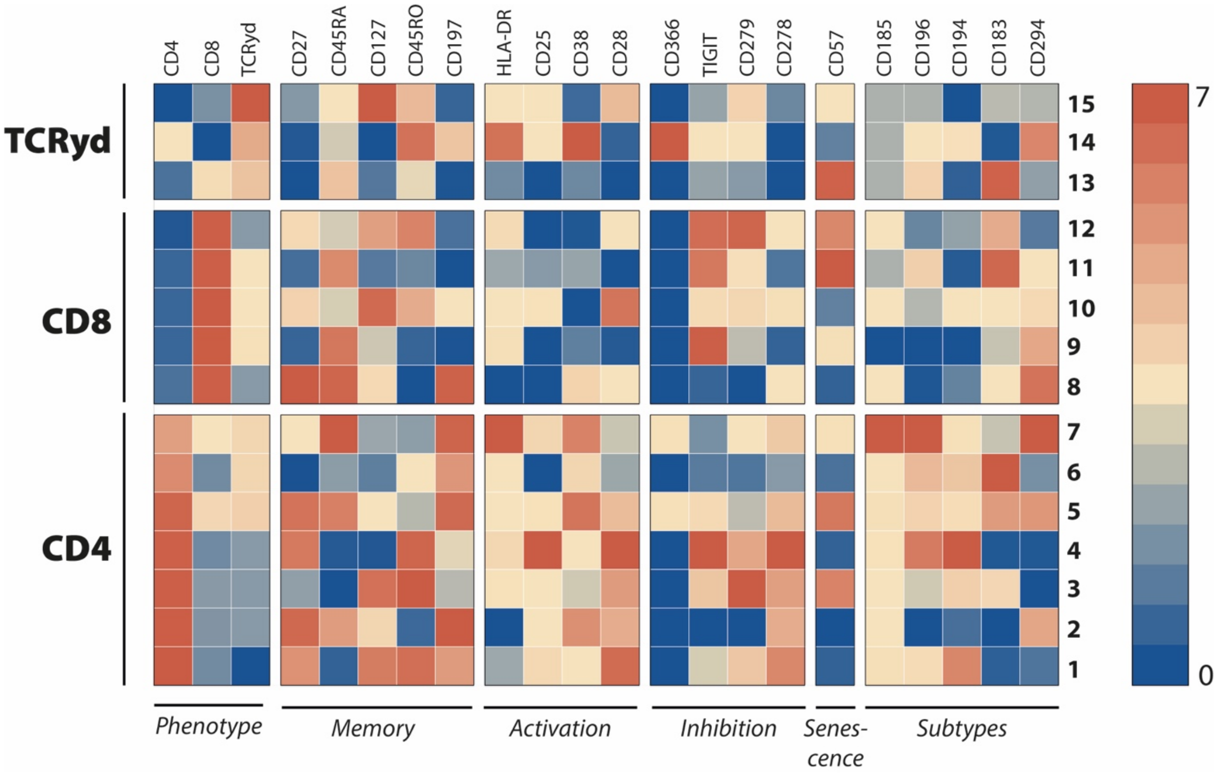


**Supplementary Figure 10. Heatmap of T-cell-based tSNE.** Relative marker expression from low (0, blue) to high (7, red) of phenotypical, memory, activation, inhibition, senescence, and Th-subtype markers on T-cells in clusters 1 to 15 at baseline.

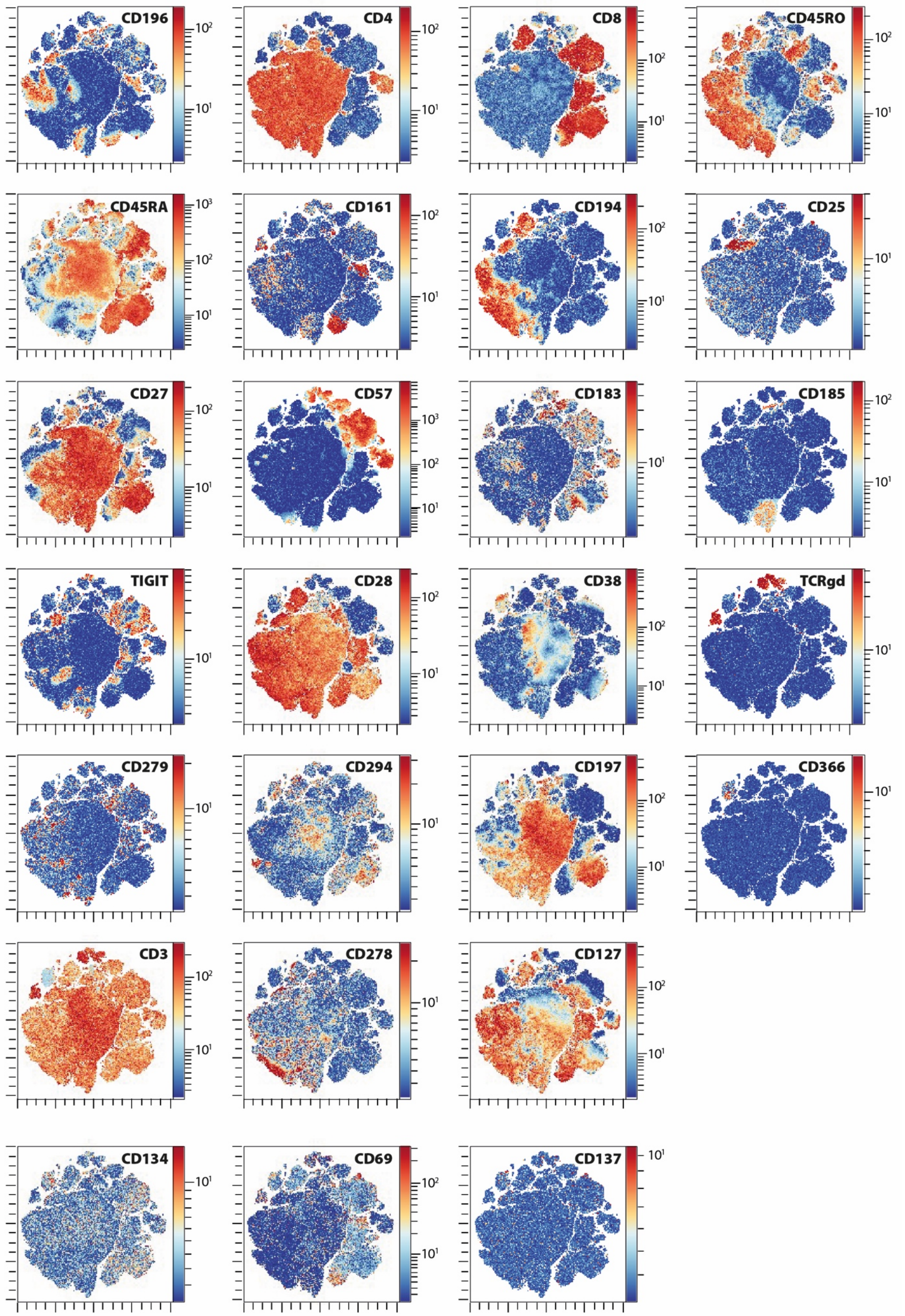


**Supplementary Figure 11. Marker expression of T-cell tSNE.** Relative marker expression of all phenotypical markers included in the optSNE on T-cells at baseline.

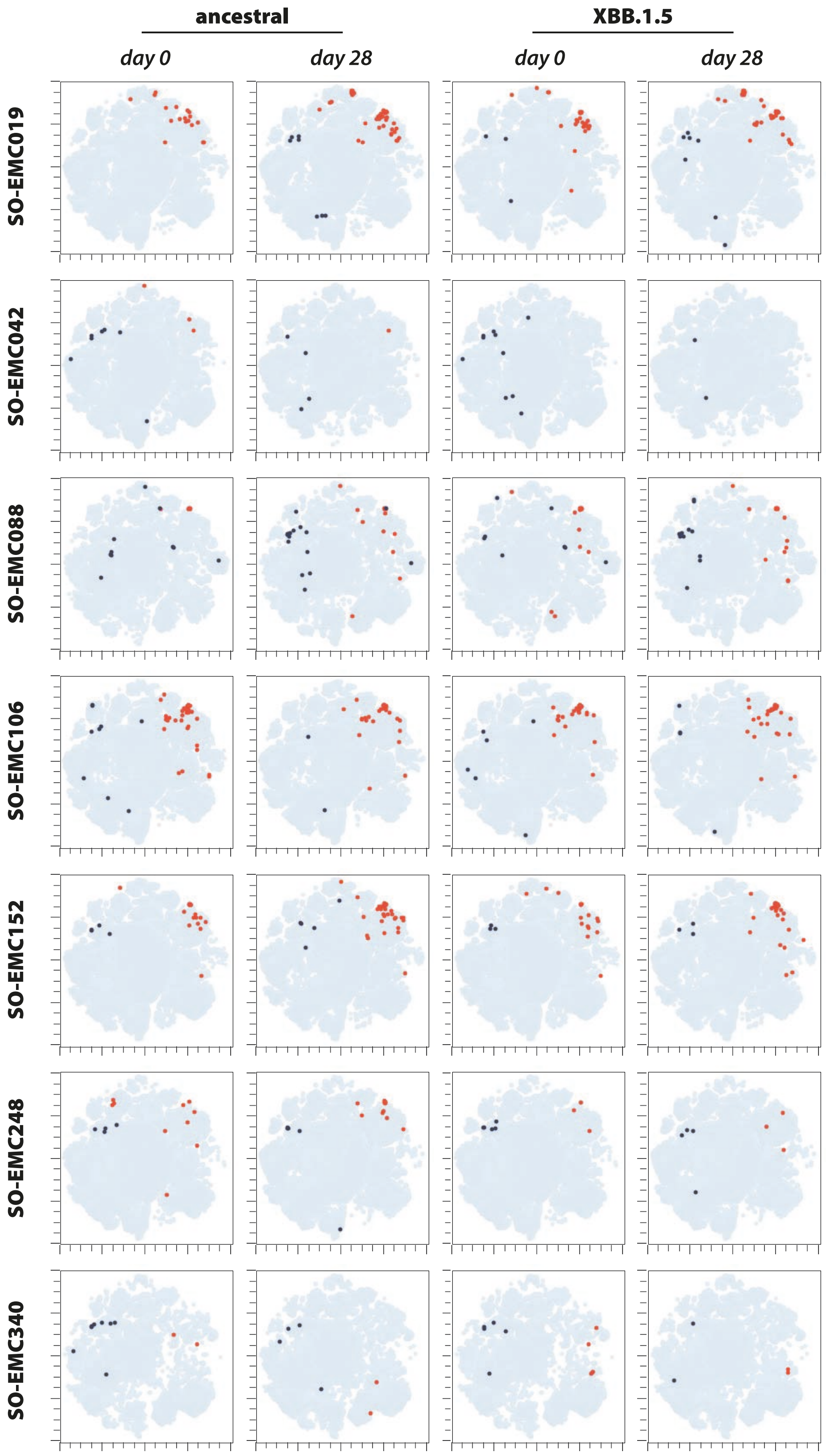


**Supplementary Figure 12. Individual AIM overlays.** Individual overlays of AIM-positive memory CD4+ (CD137+CD69+; blue) and memory CD8+ (CD137+CD69+; red) on a T-cell phenotype tSNE after stimulation with overlapping S representing ancestral SARS-CoV-2 or Omicron XBB.1.5.

### Supplementary Tables

**Supplementary Table 1.** Baseline characteristics of randomly selected participants for in-depth serological profiling

| | **Total**<br>n = 40 | **Ad/BA.1**<br>n = 9 | **mRNA/BA.1**<br>n = 11 | **Ad/BA.5**<br>n = 12 | **mRNA/BA.5**<br>n = 8 |
|---|---|---|---|---|---|
| **Demographics** | | | | | |
| Female: [n (%)] | 27 (68%) | 6 (67%) | 9 (82%) | 6 (50%) | 7 (88%) |
| Age: [years; median (IQR)] | 53 (49-56) | 54 (53-55) | 51 (36-53) | 54 (50-56) | 54 (51-59) |
| **Comorbidities** | | | | | |
| Cardiovascular disease: | 2 (5%) | 0 (0%) | 1 (9%) | 1 (8%) | 0 (0%) |
| Pulmonary disease: | 3 (8%) | 0 (0%) | 1 (9%) | 2 (17%) | 0 (0%) |
| Diabetes Mellitus: | 0 (0%) | 0 (0%) | 0 (0%) | 0 (0%) | 0 (0%) |
| Liver disease: | 1 (3%) | 0 (0%) | 0 (0%) | 1 (8%) | 0 (0%) |
| Kidney disease: | 1 (3%) | 1 (11%) | 0 (0%) | 0 (0%) | 0 (0%) |
| **Immunology** | | | | | |
| S1-binding antibodies: [BAU/mL, GMT (95% CI)] | 2460 (1540-6360) | 3560 (1650-5380) | 7240 (2080-12300) | 1805 (1235-2420) | 3660 (2350-6710) |
| **Time between visits** | | | | | |
| Last vaccination and BL: [days, median (IQR)] | 367.0 (322.5-381.0) | 381.0 (378.0-408.0) | 379.0 (375.0-384.5) | 308.5 (300.0-329.5) | 339.0 (318.5-362.5) |
| BL and 28D: [days, median (IQR)] | 28.0 (28.0-28.0) | 28.0 (28.0-28.0) | 28.0 (28.0-31.0) | 28.0 (27.8-28.3) | 28.0 (27.0-28.5) |
| 28D and 3/6M: [days, median (IQR)] | 180.0 (160.0-219.0) | 149.5 (125.8-162.0) | 159.0 (153.0-161.0) | 236.0 (214.0-246.0) | 210.0 (197.5-223.0) |

BAU: binding arbitrary units; GMT geometrical mean titer; CI: confidence interval; BL: base line; 28D: 28 days, and 3/6M: 3 to 6 months after XBB.1.5 vaccination; IQR: interquartile range

**Supplementary Table 2.** Antibody panel for memory B-cell phenotyping

| Marker | Fluorochrome | Clone | Manufacturer | Cat. number |
|---|---|---|---|---|
| **CD3** | BUV805 | UCHT1 | BD Biosciences | 612896 |
| **CD19** | BYG710 | HIB19 | Cytek Biosciences | R7-20009 |
| **CD21** | BV711 | B-ly4 | BD Biosciences | 563163 |
| **CD27** | BYG781 | O323 | Cytek Biosciences | Custom |
| **CD38** | BV605 | HB-7 | BD Biosciences | 562665 |
| **CD71** | BV786 | M-A712 | BD Biosciences | 563768 |
| **LIVE/DEAD** | ViaDye Red | - | Cytek Biosciences | R7-60008 |
| **IgA** | PE-Vio615 | RE1014 | Miltenyi Biotec | 130-116-882 |
| **IgD** | BB700 | IA6-2 | BD Biosciences | 566538 |
| **IgG1** | PE | G17-1 | BD Biosciences | 624049 |
| **IgG2** | FITC | HP6002 | BD Biosciences | 624045 |
| **IgG2** | PE | HP6002 | BD Biosciences | 624049 |
| **IgG3** | FITC | HP6047 | BD Biosciences | 642045 |
| **IgG4** | APC | SAG4 | Cytognos | CYT-IGG4AP |
| **IgM** | Hor V450 | MHM88 | Cytek Biosciences | Custom |
| **Streptavidin** | BUV395 | - | BD Biosciences | 564176 |
| **Streptavidin** | BUV496 | - | BD Biosciences | 612961 |
| **Streptavidin** | BUV615 | - | BD Biosciences | 613013 |
| **Streptavidin** | BUV737 | - | BD Biosciences | 612775 |
| **Streptavidin** | BV421 | - | Biolegend | 405225 |
| **Streptavidin** | BV480 | - | BD Biosciences | 564876 |
| **Streptavidin** | BV650 | - | Biolegend | 405232 |

**Supplementary Table 3.** CyTOF antibody panels

| Target | Clone | Manufacturer | Isotope |
|---|---|---|---|
| **Barcoding antibodies*** | | | |
| **CD45** | HI30 | Fluidigm | 113Cd |
| **CD45** | HI30 | Fluidigm | 194Pt |
| **CD45** | HI30 | Fluidigm | 195Pt |
| **Drop-in antibodies** | | | |
| **CD134** | ACT35 | Fluidigm | 142 Nd |
| **CD226** | MBSA43 | Fluidigm | 159 Tb |
| **CD69** | FN50 | Fluidigm | 162 Dy |
| **CD279** | EH12.2H7 | Fluidigm | 165 Ho |
| **Tim-3** | F38-2E2 | Fluidigm | 169 Tm |
| **CD278** | C398.4A | Fluidigm | 175 Lu |
| **CD137** | 4B4-1 | Fluidigm | 209 Bi |
| **MDIPA antibodies** | | | |
| **CD45** | HI30 | Fluidigm | 89 Y |
| **CCR6** | G034E3 | Fluidigm | 141 Pr |
| **CD123** | 6H6 | Fluidigm | 143 Nd |
| **CD19** | HIB19 | Fluidigm | 144 Nd |
| **CD4** | RPA-T4 | Fluidigm | 145 Nd |
| **CD8a** | RPA-T8 | Fluidigm | 146 Nd |
| **CD11c** | Bu15 | Fluidigm | 147 Sm |
| **CD16** | 3G8 | Fluidigm | 148 Nd |
| **CD45RO** | UCHL1 | Fluidigm | 149 Sm |
| **CD45RA** | HI100 | Fluidigm | 150 Nd |
| **CD161** | HP-3G10 | Fluidigm | 151 Eu |
| **CCR4** | L291H4 | Fluidigm | 152 Sm |
| **CD25** | BC96 | Fluidigm | 153 Eu |
| **CD27** | O323 | Fluidigm | 154 Sm |
| **CD57** | HCD57 | Fluidigm | 155 Gd |
| **CXCR3** | G025H7 | Fluidigm | 156 Gd |
| **CXCR5** | J252D4 | Fluidigm | 158 Gd |
| **CD28** | CD28.2 | Fluidigm | 160 Gd |
| **CD38** | HB-7 | Fluidigm | 161 Dy |
| **CD56** | NCAM16.2 | Fluidigm | 163 Dy |
| **TCRgd** | B1 | Fluidigm | 164 Dy |
| **CD294** | BM16 | Fluidigm | 166 Er |
| **CCR7** | G043H7 | Fluidigm | 167 Er |
| **CD14** | 63D3 | Fluidigm | 168 Er |
| **CD3** | UCHT1 | Fluidigm | 170 Er |
| **CD20** | 2H7 | Fluidigm | 171 Yb |
| **CD66b** | G10F5 | Fluidigm | 172 Yb |
| **HLA-DR** | LN3 | Fluidigm | 173 Yb |
| **IgD** | IA6-2 | Fluidigm | 174 Yb |
| **CD127** | A019D5 | Fluidigm | 176 Yb |

***Note:** A combination of $CD45^{113Cd}$ and $CD45^{194Pt}$ was used to barcode ancestral S, $CD45^{194Pt}$ and $CD45^{195Pt}$ for XBB.1.5 S, and $CD45^{113Cd}$ and $CD45^{195Pt}$ for DMSO responses.
